# A joint spatial marked point process model for dengue and severe dengue in Medellin, Colombia

**DOI:** 10.1101/2021.07.06.21260108

**Authors:** Mabel Carabali, Alexandra M. Schmidt, Berta N. Restrepo, Jay S. Kaufman

## Abstract

The spatial distribution of surveillance-reported dengue cases and severity are usually analyzed separately, assuming independence between the spatial distribution of non-severe and severe cases. Given the availability of data for the individual geo-location of surveillance-notified dengue cases, we conducted a spatial analysis to model non-severe and severe dengue simultaneously, using a hierarchical Bayesian model. We fit a joint model to the spatial pattern formed by dengue cases as well as to the severity status of the cases. Results showed that age and socioeconomic status were associated with dengue presence, and there was evidence of clustering for overall cases but not for severity. Our findings inform decision making to address the preparedness or implementation of dengue control strategies at the local level.

**Highlights:**

- A model to jointly assess the spatial distribution of reported dengue and severity.
- We account for uncertainty in the surveillance-reported dengue while modelling severe cases.
- We assess spatial clustering of dengue and severe dengue cases in Medellin.
- Non-monotonic distribution of reported dengue cases across socioeconomic status.

## 1. BACKGROUND

Dengue is a vector-borne viral disease transmitted to humans by *Aedes* mosquitoes and an important public health problem worldwide (1-4). The clinical presentation of dengue ranges from a self-limited mild febrile illness to severe outcomes (5, 6). Although lifelong immunity can be developed for each one of the four dengue serotypes (7-9); secondary or subsequent infections from different dengue serotypes increase the risk of severe dengue. Severe dengue is a potentially fatal complication with case fatality rates (CFR) up to 20%. However, under adequate access to health care, diagnosis and management, dengue CFR could be as low as 1% (3, 10, 11).

Colombia is one of the Latin American countries with the highest burden of dengue (3, 4) and within Colombia, dengue burden concentrates geographically in 50 of the 778 municipalities that routinely report dengue cases (1, 2, 12, 13). Given the spatial heterogeneity of dengue distribution, including its concentration in low socioeconomic settings, and the limitations of current dengue control strategies (14-18), it is important to investigate the spatial distribution of dengue cases. For instance, it is necessary to understand how the observed case-specific characteristics, in addition to area level covariates, are associated with the distribution of overall dengue cases. In addition, the analysis of severe cases is usually performed separately from the analysis of overall reported cases, assuming independence between overall presence of dengue and the presence of severity, often ignoring the potential underreporting associated with the use of surveillance data (4, 15, 19-25). Assuming independence between non-severe and severe cases distribution potentially leads to underestimation of the severity and the uncertainty associated to the individual factors related to severe cases (6, 14, 18, 25). Moreover, while analyzing severe cases, it is important to identify whether the distribution of severe cases follows a different spatial distribution from that of the overall notified dengue cases. However, such analyses are rare and limited because they are: i) often constrained by data availability, ii) mainly conducted using aggregated census-area level data only, iii) often lacking proper adjustment of neighboring areas, and iv) usually computationally intensive (4, 12, 15, 16, 19-23).

Given the availability of individual dengue case locations (exact longitude and latitude), and to identify high-risk dengue areas while modelling *simultaneously* non-severe and severe cases, we conducted a single joint spatial marked-point-processes model of notified dengue cases in Medellin, Colombia. We were motivated by the advantage of using individual level location and area level information to identify spatial patterns for clustering areas while properly accounting for spatial autocorrelation (20, 21). Hence, the main purpose of this study is to present the methodology and to estimate quantitatively the contribution of area- and the observed case-specific characteristics while jointly analyzing the spatial distribution of notified (i.e., surveillance data) dengue and severe dengue in Medellin, Colombia.

## 2. METHODS

### 2.1. Study site

Medellin is the second largest city in Colombia with 2.6 million inhabitants (26). Annual dengue incidence ranged between 161 and 745 cases per 100,000 inhabitants over the last 10 years (1) and is consistently included on the top five dengue-reporting cities since 1998 (2). Medellin’s urban area is composed of 269 neighborhoods, including 20 institutional units such as university campuses, jail facilities and military compounds, distributed over 110 km^2^. Medellin’s altitude ranges from 1,460 to 3,200 meters, the annual average temperature is 24°C, and it has two rainy seasons (April and October). Although 50% of the city is classified as low socioeconomic status (SES), 98% of the city has access to potable water. The distribution of health coverage of the population is 70% contributory (employees or self-employees), 25% government subsidized, and 4% uninsured (26).

### 2.2. Data description

To illustrate the proposed methodology, we use an available dataset including observations of individual location (exact longitude and latitude) of all notified dengue cases in Medellin in 2013 (n=1,793). Dengue notification in Colombia is mandatory and cases are individually registered in the national surveillance system (SIVIGILA), using the locally validated and standardized codes 210 and 220 for dengue and severe dengue, respectively (27). We chose to use this dataset given the availability and to avoid the potential threat of misclassification with other arboviruses when using data of notified cases from 2014 and onwards, when Chikungunya and zika were introduced in the country (13, 28).

#### Individual level covariates

Each row of the dataset included individual sociodemographic and clinical information for each notified case, including sex, age categorized as under 15 years; 15 to 34 years; 33 to 54 years; and over 55 years and for aggregated analysis as proportion of cases under 20 years of age and proportion of cases over 20 years of age, residential and work/study addresses, date of notification, date of symptoms’ onset, severity status, insurance scheme (subsidized vs contributory schemes) (29), and neighborhood of residence, all collected routinely in SIVIGILA’s notification form (27).

#### Area level covariates

The neighborhood’s population and socioeconomic status index (SES) were obtained through the office of development and planning at the local ministry of health and the Colombian Administrative Department of Demographic Survey (DANE) (26, 30). Entomological information, including the Breteau Index (IB) which is usually categorized as low, medium or high (27), was used to determine the neighborhood specific level of *Aedes* infestation and obtained from Medellin’s local secretary of health. However, according to the entomological information reported for the year of study, there were no neighborhoods with high Breteau Index for that year’s entomological survey.

### 2.3. Study design

We performed a spatial analysis using a single joint spatial marked point process model, to simultaneously estimate the underlying process leading to the spatial patterns of overall and severe dengue cases (22, 31).

#### 2.3.1. Spatial point process model

A spatial point process assesses the distribution of the individual location of an outcome, over a spatial region (22, 23). Here, the individual spatial location (exact longitude and latitude) of an outcome is denominated by a point pattern (22, 23, 32). There are several other proposed models used to assess the point pattern distribution of dengue, including the analysis of disease transmission using agent-based models (33), and analysis using space-time kernel density estimation (34, 35). Here, we propose a model-based approach wherein the logarithm of the intensity of notified dengue cases across Medellin is modelled through a latent Gaussian random field (22, 31, 32).

Specifically, we proposed a Log-Gaussian Cox process which given the nature of the point process follows a Poisson distribution (22, 23, 32, 36). As the likelihood function of a Log-Gaussian Cox process involves an integral that does not have an analytical solution, we used the neighborhood structure of Medellin to approximate this integral. See for example Pinto Jr. *et al*., (36) for details about this approximation. To identify whether there is an underlying mechanism leading to a different spatial distribution of severe cases, we considered the presence of severity as an individual characteristic of each case and attributed it as a “mark” of the individual point. Since the presence of severity is conditional on being a case, we cannot assume independence between overall notified cases and severe cases. Therefore, the number of severe cases, conditioned on the total number of reported cases in each neighborhood, is assumed to follow a Binomial distribution. For this Binomial distribution, the probability of presence of severity is described by the fixed effects of the observed severe cases characteristics and an area latent spatial effect, which is assumed to be proportional to the one used in the mean of the Poisson distribution for overall dengue cases. Here, the proposed approach has the advantage of *simultaneously* assessing the spatial distribution of overall dengue cases and severe cases, by considering the spatial autocorrelation between spatial units. In other words, as the relative risk of cases and the probability of severe dengue share a common component, the joint approach allows us to learn about this underlying spatial process. Another advantage includes accounting for the uncertainty associated with the reported number of dengue cases in the surveillance-based data (4, 19, 25, 31, 32) (Supplementary Material).

#### 2.3.2. Model description

To fit a joint spatial marked-point-processes model we first constructed a model for each latent random field, one for the “pattern”: overall cases, and another for the “marks”: severe cases (32), which are specified as follows:

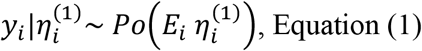

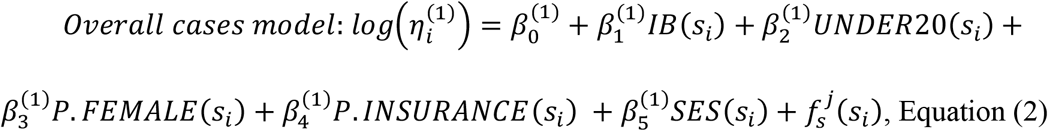

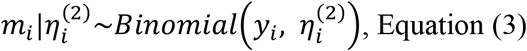

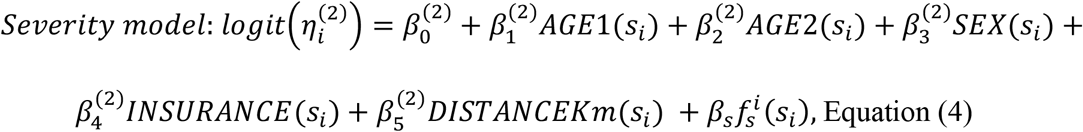

Here, we assume that *y*_*i*_, the total number of dengue cases observed in each neighborhood *i*, follows a Poisson distribution with mean 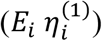, where *E*_*i*_ is the expected count of cases in neighborhood *i*, obtained via indirect standardization using the city’s disease rate (37) and 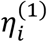 is the Standardized Rate Ratio (SRR) for neighborhood *i*. Following equation (2), the logarithm of SRR is decomposed as the sum of areal effects and a spatially structured random effects 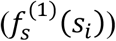 modelled following the Besag specification (38, 39). The component 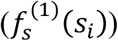 follows, *a priori*, a Gaussian Markov Random Field (GMRF) and accounts for the spatially structured effect for the pattern, which reflects the spatial autocorrelation (neighboring structure or vicinity) in the latent field that is not explained by the covariates (i.e., fixed effects) (20, 32, 40). Other components of the overall cases model included 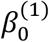 which is the pattern’s intercept and as fixed effects for the pattern of overall dengue cases we included the following neighborhood level covariates with their corresponding *β*^(1)^ coefficients including the observed cases-characteristics such as the proportion of dengue cases under 20 years of age (*UNDER20*_*i*_); the proportion of female dengue cases (*P. FEMALE*_*i*_), the proportion of cases with contributory scheme cases (*P*.*INSURANCE*), and area-level characteristics data obtained from entomological survey Breteau Index (*IB*_*i*_) categorized as low or medium given that there were no neighborhoods with high IB; the socioeconomic status level (*SES*_*i*_), a categorical variable with three levels (low SES level, medium SES level, and high SES level) obtained from DANE(30).

For the analysis of the severity “marks”, in equation (3) *m*_*i*_ is the number of severe cases in each neighborhood *i* and follows a Binomial distribution, 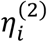 is the probability of cases being severe case in neighborhood *i*, and *y*_*i*_ is the total number of dengue cases observed in each neighborhood *i*. The logit 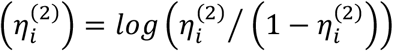 is the random field for the marks (severity) and the exponentiated *β*^(2)^ coefficients are the odds ratio (OR) for severity: 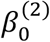 is the marks’ intercept and the fixed effects covariates for the severity included the proportion of severe cases under 20 years *AGE*1(*s*_*i*_) and over 20 years *AGE*2(*s*_*i*_); the proportion of female severe cases: *SEX*(*s*_*i*_); the proportion of severe cases using contributory scheme: *INSURANCE*(*s*_*ij*_); and the minimum distance between severe cases per neighborhood *DISTANCEKm*(*s*_*ij*_), which is the standardized nearest-neighbor (Euclidean) distance (km) between severe cases in each neighborhood. The logit of the probability of severity is decomposed as the sum of the fixed effects described above and a latent spatial effect, 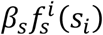 that is proportional to the spatial latent effect in the log relative risk of overall dengue cases 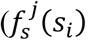, in equation (2)). The component 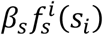 in equation (4) represents a single (common) random field, and includes a coefficient *β*_*s*_ that makes the structured spatial effect for the severity 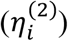 proportional to the spatial effect of the pattern 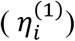 (32); which is justified given that being a severe case is conditional on being a case in the first place. Therefore, it is expected that these two processes share common characteristics after accounting for the different effects of the available covariates.

#### 2.3.3. Data analysis

We calculated the respective descriptive statistics, and continuous estimates were presented as mean and standard deviation (SD) or as median and Interquartile Range (IQR), while categorical variables were presented as proportions. To inspect the observed distribution of cases, we plot the kernel density of the individual overall and severe dengue cases using a 5 km bandwidth (41).

The proposed joint spatial marked-point-processes model represents the two outcomes (overall reported dengue cases and severity) *simultaneously* in a hierarchical mixed-effects Bayesian model. The overall disease pattern and the severity marks constitute a matrix outcome of two link functions (i.e., Poisson for overall dengue cases, and Binomial for severity); the parameters for the Poisson and Binomial distributions were jointly analyzed in relation to the vector of the sociodemographic covariates described above (32). For the overall dengue pattern, we estimated the crude and adjusted Standardized Rate Ratio (SRR). For the severity marks, we estimated the odds ratio (OR), the respective probability of severity, and the overall and neighborhood-specific Relative Risk (RR) of severity. We assigned non-informative priors for the precision parameters of the random effects. The posterior distributions of the parameters and respective 95% Credible Intervals (95% Cr.Int) were estimated via Integrated Nested Laplace Approximation (INLA) (32, 40, 42). The variables included in the full models: equation (2) and (4), were chosen after consideration of different set of relevant covariates. Model selection was performed through the Deviation Information Criterion (DIC) (20, 22, 32). All analyses were fitted using R-INLA (R Core Team (2019); R Studio version 3.3.3) (23, 42, 43). We followed the REporting of studies Conducted using Observational Routinely-collected health Data (RECORD) statement guideline (44) (Supplementary Material).

##### Sensitivity analysis

To test the performance of our proposed method we also fitted the proposed joint model using separate spatial structures for patterns and marks, evaluated different structured spatial effects by including an independent unstructured random effect for the pattern (overall cases) and for the marks (severe cases), denoted *u*(*s*_*i*_) and *v*(*s*_*i*_) in equations (2) and (4), respectively, and which were parameterized using the Besag-York-Mollié (BYM) specification (38). We also explored the BYM2 which is a reparameterization of the BYM and included a mixing parameter (Phi) which help with the interpretation as an indicator of spatial dependency(45) (Supplementary Material).

##### Ethics statement

This study analyzed secondary data without identifying information, and therefore, did not require informed consent. The protocol was reviewed and approved by the Institutional Review Board (Study No. A02-E05-18A) and by the ethics committee of the Secretary of Health of Medellin, Colombia.

## 3. RESULTS

In 2013, there were 1,793 dengue cases reported in Medellin. In total, 1,719 (96%) were geocoded and were used for this analysis. There were 247 (14.4%) severe cases. Median age was 28 years (IQR=16 - 45) for overall dengue cases and 29 years (IQR=17 - 49) for severe cases. A descriptive analysis of notified cases and neighborhood characteristics is presented in **Table 1**.

**Table 1.** Individual and Neighborhood Characteristics of Dengue Cases Reported in Medellin, 2013.

|  | **Overall cases** | **Severe cases** |
| --- | --- | --- |
| **Individual Characteristics** | **n (%)** | **n(%)** |
| Complete cases | 1,719 | 247 (14.4) |
| **Age, median (IQR)** | 28 (16, 45) | 29 (17, 49) |
| <15 years | 370 (21.5) | 49 (19.8) |
| 15-34 years | 710 (41.3) | 100 (40.5) |
| 35-54 years | 424 (24.7) | 58 (23.5) |
| >55 Years | 215 (12.5) | 40 (16.2) |
| **Sex (Male)** | 884 (51.4) | 130 (52.6) |
| **Insurance** |  |  |
| Subsidized | 567 (33) | 90 (36.4) |
| Contributory | 1,152 (67) | 157 (63.6) |
| **DENV Classification** |  |  |
| Severe Dengue | 247 (14.4) | - |
| Hospitalization | 384 (22.4) | 185 (74.9) |
| **Distance between cases in Km, median (IQR)** | 5.15 (0.0, 12.1) | 0.2 (0.1, 0.3) |
| **Neighborhood Characteristics** | **n (%)** | **n (%)** |
| Low SES Level | 641 (37.3) | 90 (36.4) |
| Medium SES Level | 885 (51.5) | 128 (51.8) |
| High SES Level | 193 (11.2) | 29 (11.7) |
| Breteau Index (Low) | 999 (58.2) | 132 (53.4) |
| Breteau Index (Medium) | 717(41.8) | 115 (46.6) |
| Proportion of Female cases*, median (IQR) | 50.0 (40.0, 60.0) | 50.0 (37.5, 56.0) |
| Proportion of cases* <20 years old, median (IQR) | 32.0 (20.0, 44.8) | 32.0 (20.0, 43.8) |

*\*Among the total reported cases per neighborhood.*

The overall crude rate for reported dengue was 78 cases per 100,000 inhabitants. The median number of cases per neighborhood was four (IQR= 1-10; range= 0-57). The mean crude SRR was 1.3, standard deviation (SD: 2.4; range= 0-20.6). The median number of severe cases per neighborhood was one (IQR=0-2; range= 0, 10). There was an apparent concentration of both dengue and severe dengue cases on the northeastern neighborhoods that was observed by the crude distribution of geocoded cases (**Figure 1**) and the unadjusted (i.e.: without accounting for population size) estimated density of cases (Supplemental Material).

**Figure 1.**
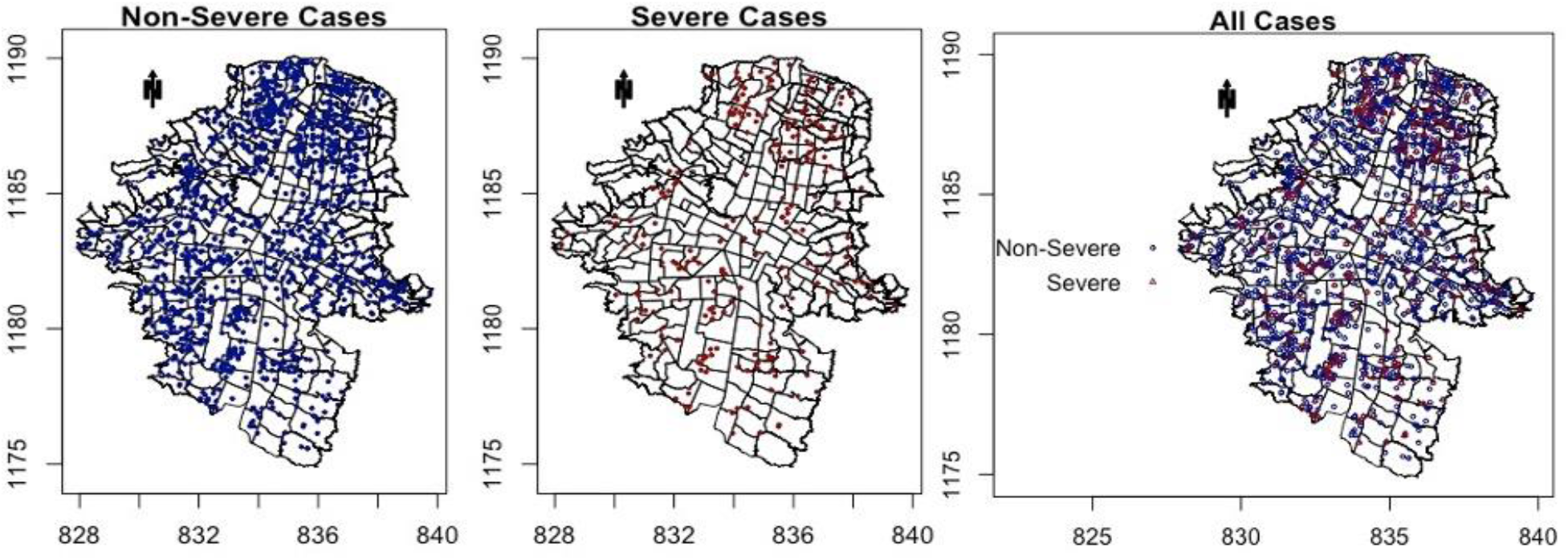
Distribution of dengue cases notified in Medellin in 2013. Plot of observed overall and severe dengue cases.

The spatial distribution of the unadjusted SRR for overall dengue cases indicated the presence of dengue in the entire city with some concentration of dengue cases among neighborhoods in the central and the North-Eastern regions of the city. Likewise, compared to the overall odds of severity in the entire city, the distribution of severe cases indicated increased odds of severity among Southern and Eastern neighborhoods of the city (**Figure 2**).

**Figure 2.**
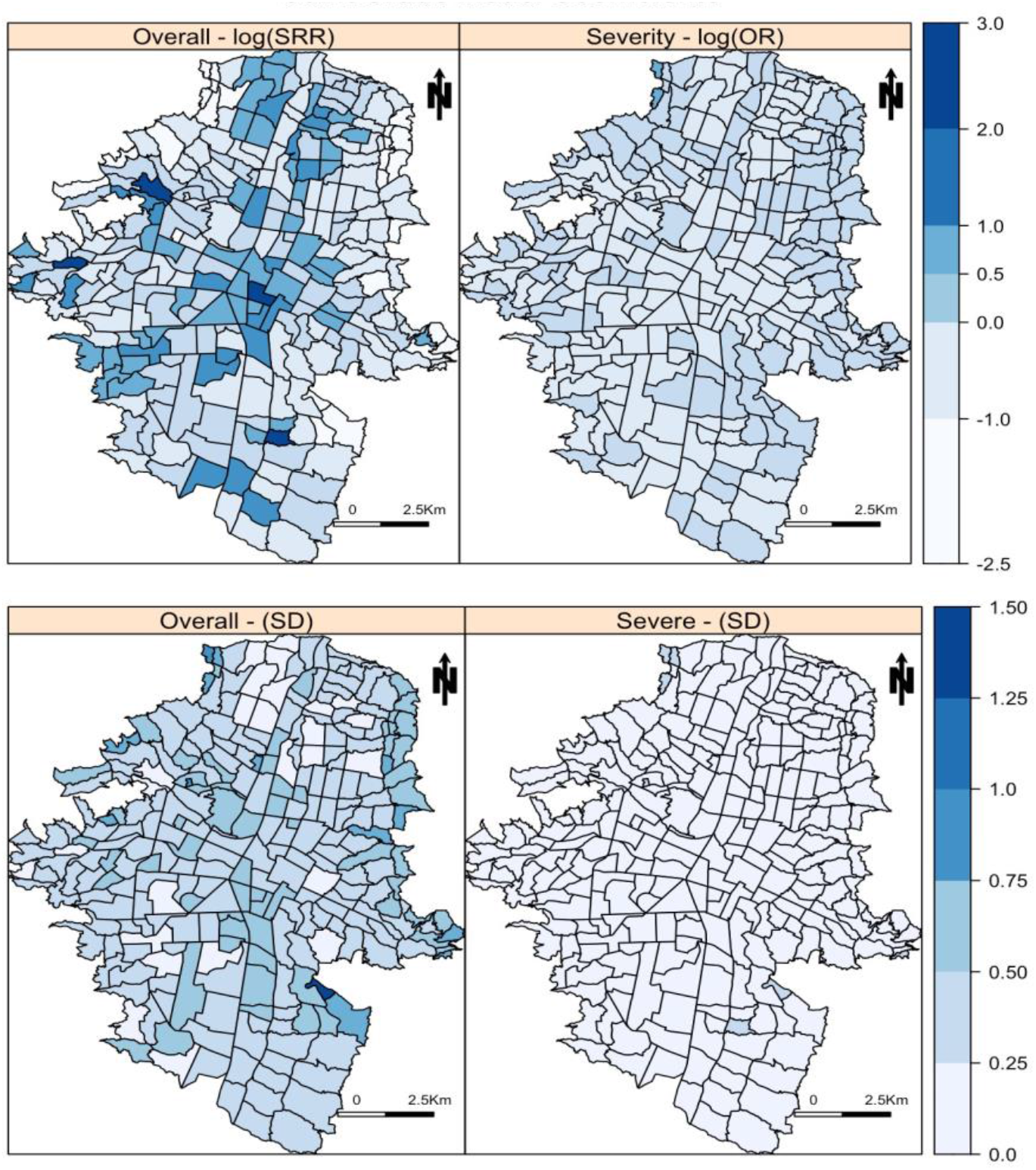
Spatial distribution of overall and severe dengue cases in Medellin, 2013. (Top) Neighborhood specific crude Standardized Rate Ratios (SRR) and standard deviation (SD) for overall dengue cases. (Bottom) Neighborhood specific crude Odds Ratios (OR) and standard deviation (SD) for severe dengue cases. Note: Scales are different given the magnitude of the estimates.

The fully adjusted proportional joint model showed that given the presence of dengue cases the median adjusted probability of severity per neighborhood was 18.7% (95% Cr.Int=5.3%, 55.7%). Overall dengue rates increased with every 10% increase in the proportion of cases under 20 years old per neighborhood (SRR=1.10; 95% Cr.Int=1.04, 1.16), the proportion of female cases (SRR=1.07; 95% Cr.Int=1.03, 1.12), and cases with contributory insurance scheme (SRR=1.07; 95% Cr.Int=1.02, 1.12). Just over half of reported cases were from neighborhoods with medium SES levels, and compared to these, dengue rates among neighborhoods in the Low SES level were on average 26% lower (SRR=0.74; 95% Cr.Int=0.56, 0.98) and rates among neighborhoods with high SES level were on average 31% lower (SRR=0.69; 95% Cr.Int=0.48, 1.01). There were no neighborhoods with a high Breteau Index (i.e., high *Aedes* presence) and the comparisons were made between low (reference) and medium Breteau Index levels. Compared to neighborhoods with low Breteau Index (i.e., low *Aedes* presence), neighborhoods with a medium level of Breteau Index had slightly higher rates of dengue cases (SRR=1.06; 95% Cr.Int=0.83, 1.36) but the credible intervals covered the null value. For severity, the posterior distribution of the proportion of severe female cases, severe cases under and over 20 years of age, and severe cases with contributory insurance scheme coefficients showed high posterior uncertainty with 95% credible intervals including the null value. (**Table 2**).

**Table 2.** Posterior mean of the Standardized Rate Ratio (SRR), the Odds Ratio (OR) and 95% Credible Intervals (95% Cr.Int) for covariates (fixed effects) in the joint proportional model for dengue cases in Medellin, 2013 (DIC=1578).

| Fixed Effects Covariates | Standardized Rate Ratio or Odds Ratio and 95% Credible Intervals |  |  |
| --- | --- | --- | --- |
|  | SRR | 2.5% | 97.5% |
| **Overall Cases (Area Level Covariates)** |  |  |  |
| Proportion of Female Cases | 1.07 | 1.03 | 1.12 |
| Proportion of cases <20 years old | 1.1 | 1.04 | 1.16 |
| Proportion of Contributory scheme | 1.07 | 1.02 | 1.12 |
| Low Breteau Index | Ref. | - | - |
| Medium Breteau Index | 1.06 | 0.83 | 1.36 |
| Medium SES Level | Ref. | - | - |
| Low SES Level | 0.74 | 0.56 | 0.98 |
| High SES Level | 0.69 | 0.48 | 1.01 |
| **Severe Cases** | **OR** | **2.5%** | **97.5%** |
| Proportion of Female Cases | 0.97 | 0.93 | 1.01 |
| Proportion of cases <20 years old | 1.04 | 0.88 | 1.20 |
| Proportion of cases >20 years old | 1.06 | 0.90 | 1.23 |
| Proportion of cases with Contributory Insurance | 0.99 | 0.95 | 1.03 |
| Median distance between severe cases (Km) | 0.99 | 0.98 | 1.00 |
| **Spatial Effect ($\beta_s$)** | 0.78 | 0.65 | 0.93 |

**Area Level Covariates:** **Proportion of Female Cases:** indicates every 10% increase in the proportion of female cases reported per neighborhood; **Proportion of cases <20 years old:** indicates every 10% increase in the proportion of reported cases <20 years old per neighborhood. **Proportion of Contributory Scheme Cases:** indicates every 10% increase in the proportion of cases with contributory scheme insurance reported per neighborhood; **Breteau Index:** Comparing the Low Breteau Index level (Reference group) to Medium Breteau Index level; **SES Level:** Comparing cases in the Medium SES (Reference group) to Cases in the Low and High SES levels. **Spatial Effect** = $\exp(\beta_s \text{ Coefficient from } g_s^j(s_{ij}))$ in Equation 4.

After adjusting for other covariates and comparing to the overall rate of dengue in the city, the spatially structured effect indicating the residual spatial autocorrelation not explained by the fixed effects, showed a widespread distribution of cases with some concentration in central and Northern parts of the city. For severity marks, the residual spatial effect showed a homogeneous distribution of severe cases without indication of concentration of cases in any particular neighborhood (**Figure 3**).

**Figure 3.**
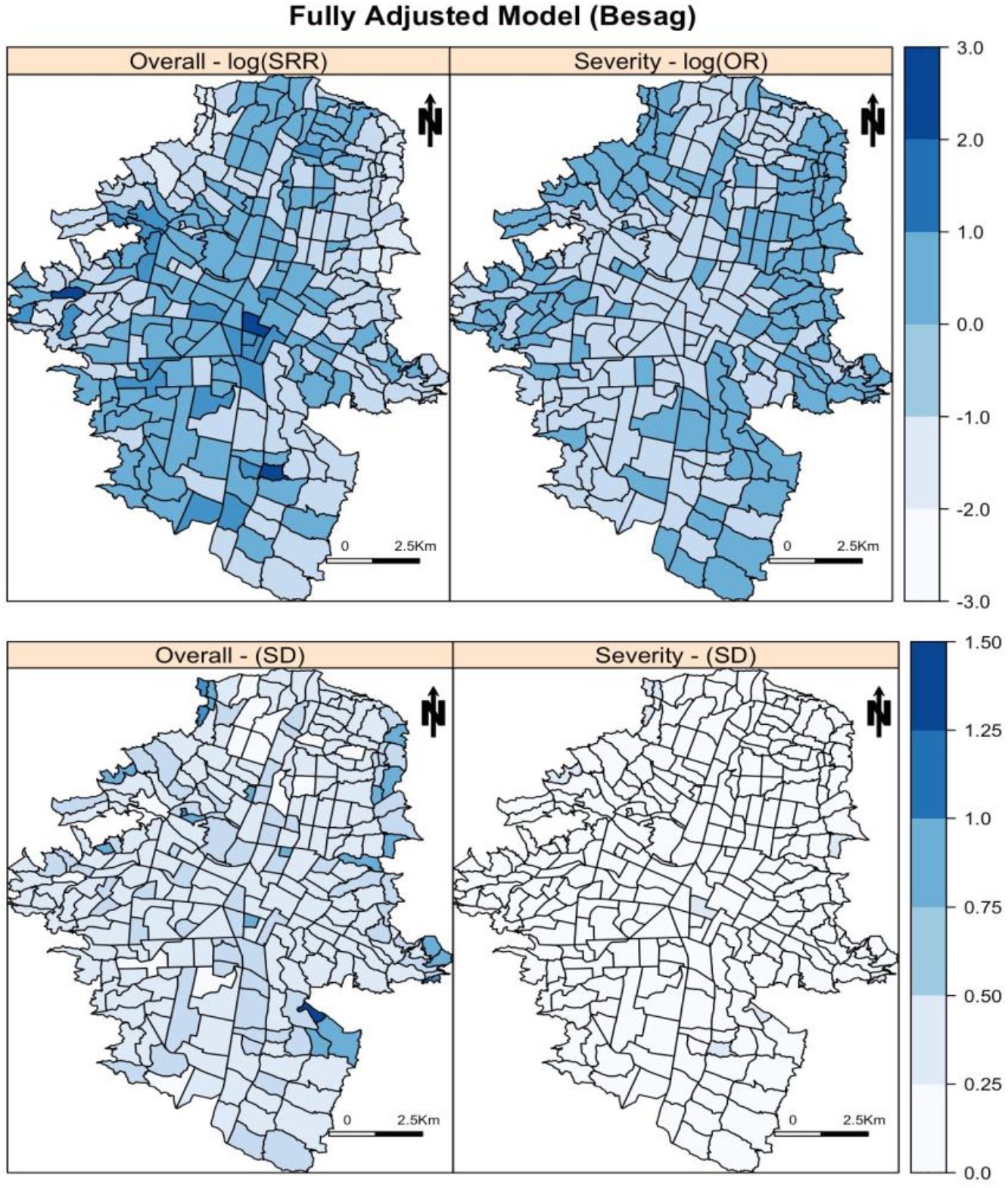
Estimated common spatial trend for overall dengue and Severe dengue cases in Medellin, 2013. (Top) Neighborhood specific residual (Random Effects) Standardized Rate Ratio (nSRR) and Standard Deviation (SD). (Bottom) Neighborhood specific residual (Random Effects) Odds Ratio (nOR) and Standard Deviation (SD).

The spatial model estimated the marginal variance of the spatial effect (derived from the precision parameter for the spatial correlation) and a Beta coefficient *β*_*s*_ from equation (4), to indicate the proportionality between the spatial structure of overall cases and severe cases. The joint model showed moderate marginal variance of the spatial effect for dengue distribution (Variance= 3.02; 95%Cr.Int= 2.34, 4.00). The Beta coefficient *β*_*s*_ for the spatial effect of severe cases in equation (4) indicated that after accounting for the other covariates in the model, the distribution of overall dengue cases and the distribution of severe dengue shares the latent spatial effect but in smaller magnitude (OR= 0.78; 95% Cr.Int=0.65, 0.93).

As a sensitivity analysis we fit the joint model using separate spatial structures for patterns and marks. The results from the mean of the posterior distribution for the fixed effects were similar to the main results presented here but showed higher posterior uncertainty with wider credible intervals, mostly for SES and age for the overall dengue distribution and for all the covariates for severity. The DIC for the model using separated spatial structure was 1590 the model with the joint proportional structure was 1578, suggesting that the model that assumes that the log relative risk of dengue cases and the logit of odds ratio of severe cases share a spatial effect fits the data better. (Supplementary Material). The hyperparameters for the spatial effect in the model with separated structures showed more variability in the marginal variance for the spatial structure for overall cases (Variance=0.60; 95%Cr. Int= 0.43, 0.82) than for the distribution of severe cases (Variance =0.02; 95%Cr. Int= 0.005, 0.33). Compared to the joint model, using only independent spatial structures, the distribution of severe cases become independent of the distribution of non-severe cases. Since the assessment of the spatial distribution of severe cases in the separate model does not account for the spatial distribution of overall cases (i.e., *there is no borrow of* strength across the overall spatial distribution of dengue cases), the estimates of the fixed effects and the hyperparameters for severity are less precise, with wider confidence intervals for the fixed effects ORs and the marginal variance. This suggests that the use of the joint model with the proportional spatial structure that borrows the strength across the spatial distribution of overall and severe cases is preferred. Moreover, epidemiologically, it is relevant and plausible given that dengue and severe dengue are transmitted in the same way, and severity is a potential clinical change in the status of a dengue case.

The BYM model estimated two hyperparameters for the precision of the structured spatial component and another for the unstructured component and BYM2 models estimated the marginal variance (derived from the precision parameter for the spatial correlation) and an indicator of spatial dependency (Phi). The coefficients for the fixed effects models using BYM and BYM2 parameterizations were similar to estimates using the Besag parameterization and the estimated posterior distribution of the spatial common trend were similar but the marginal variance for the structured spatial component was not identifiable with the BYM parameterization. The joint models using the BYM2 structure showed smaller marginal variance of the spatial effect (Variance=0.69; 95%Cr. Int= 0.54, 0.86) and a 22% tendency of spatial dependency for the distribution of overall cases (Phi= 0.22; 95%Cr.Int = 0.03, 0.57). The Beta coefficient (*β*_*s*_) in all cases indicates proportionality with the overall distribution of cases in smaller magnitude (Supplementary Material).

## 4. DISCUSSION

We presented an analysis of a joint spatial marked point processes model on routinely collected dengue data. Our proposed approach shows the possibility of *simultaneously* estimating the distribution of overall dengue cases and the distribution of severity, which given the shared common spatial component, allows the assessment of any underlying spatial process. The proposed approach also contributes to account for the uncertainty associated to the reporting of dengue cases in surveillance-based data, allows for spatial autocorrelation, and uses case-specific observed sociodemographic covariates to explain both outcomes: dengue and severe dengue.

### 4.1. Dengue discussion

Colombia is an endemic country and Medellin is one of the municipalities consistently reporting a high burden of cases during the last decade (1, 2, 27). Our study shows that during 2013, dengue was present in the entire city, with concentration at the Northeastern neighborhoods, which are known for being densely populated areas (18, 26, 46). The concentration of cases in the Eastern region of the city has been previously explored in the context of serological surveys (47) and among children attending different schools in the city (18). However, previous approaches did not include latent spatial structured effects that account for the neighboring structure after adjusting for available covariates. Also, previous approaches either used only census aggregated data to fit overall dengue cases and severity separately, fitted fixed effects for the spatial structure or modeled separately the spatial effects and the contribution of case-specific observed sociodemographic covariates (14, 18, 34-36, 46, 47).

In our study, there were no neighborhoods with high *Aedes* presence determined by the Breteau Index and there was no association between the SRR and the presence of medium level Breteau Index. Although the Breteau Index is considered a useful indicator of *Aedes* infestation, there is conflicting evidence about the concordance with presence of dengue cases (1, 15). This could arise in our data because entomological information was collected at regular intervals throughout the year in different neighborhoods and households (18, 27, 46). The value of entomological indexes changes over time, but the timing of exposure assessment and incident cases may not be aligned (48-50).

Although the proportion of female cases was associated with a slight increased rate in the overall distribution of cases, being female was not associated with severity in our study. Increased proportion of female dengue cases has been also reported in Medellin previously (18). However, associations between sex, dengue and dengue severity have been inconsistent in the literature (11, 46, 47). Age, specifically the proportion of people under 20 years of age, was associated with increased rates of overall dengue cases across neighborhoods and an increased OR for severity was observed among people over 20 years of age in the proportional model and among individuals 55 years old in the sensitivity analysis. These findings could be associated with a high seroprevalence of dengue in the city and a limited presence of secondary infections (2, 11, 46). In Medellin, the overall dengue seroprevalence was estimated at 61%, with a mean age of 30 years among dengue seropositive cases. The overall seroconversion rates were estimated to increase with age, with the highest seroconversion rate (17.9 per 1,000 people) observed among subjects between 31 and 40 years of age (47). Likewise, among school children under 19 years old, a trend of increased dengue seroprevalence and seroconversion with age has been reported (18). However, it is also possible that the observed trend of severity by age could be related to comorbidities in older patients and the possibility of secondary infections in people over 55 years old (6, 9, 11). These characteristics have been described in other Colombian municipalities and in other Latin-American contexts (1, 2, 11, 14, 16, 24, 28, 46); and may contribute to an understanding of the age-related findings in this study.

In Colombia, the health coverage under the contributory system corresponds to employed individuals or people with capacity to pay for their health system coverage (affiliated to a private insurance plan or out-of-pocket) and the subsidized system corresponds to individuals for whom the state pays for health coverage. Health insurance here was modeled as a proxy for socioeconomic status at the individual level (30, 51, 52). In our study, most of the notified cases were from the contributory system and contributory insurance was associated to the overall dengue rates but not to severity (1, 2, 51-53). Using the health insurance as a proxy for individual socioeconomic status could be challenging but modeling simultaneously area level-socioeconomic characteristics and individual-level socioeconomic status or their proxy addresses this issue, at least partially. For example, a person earning a minimum wage could indeed have a contributory insurance, which could be understood as a person in a “better standing”. However, since a person earning a minimum would less likely live in a high SES neighborhood, the regression-based adjustment for the two variables in our joint model is considered pertinent. According to the SES level of the neighborhood of residency, findings from the joint model suggests a non-monotonic distribution of cases across SES levels, with fewer cases at low and high SES levels. There were fewer cases among neighborhoods at the lowest SES level, which could be attributed to limited access (physical and financial) to health care, compared to people living in neighborhoods with medium- or higher SES levels and or health seeking behavior (1, 2, 51-53). Although the rate of dengue cases seems to decrease in neighborhoods at the high SES level, the lack of precision (i.e., high uncertainty of the posterior distribution) of the estimates could be attributed to the small number of cases in this stratum (n=193 cases). Nonetheless, reporting bias and spatial confounding associated to the SES level and health seeking behavior by affiliation to health insurance could not be completely ruled out (54).

### 4.2. Implications of routinely collected data

We used passive surveillance data, which implies a potential risk of under reporting and measurement error (17, 19, 25, 27, 52, 55). Notification depends on health seeking behavior, which in turn depends on presence and severity of symptoms and access to health care (insurance scheme, availability of health care facility, etc.) that altogether could also depend on other socioeconomic factors (15, 52, 55). Therefore, the findings from this analysis should be restricted to the subset of notified cases. For this analysis we worked closely with the municipality’s secretary of health, which is considered one of the strongest surveillance systems in the country and for which dengue is a disease of mandatory notification (18, 27, 47). The diagnostic system in place, including serological and clinical confirmation, and the use of data from before the introduction of other arboviruses (i.e., chikungunya and Zika), decreased the risk of misclassification of the outcome but did not rule it out completely. To illustrate the proposed methodology, we used the available information of cases reported in 2013. Therefore, analyses using more recent data are encouraged, although given the endemicity of dengue in the study site, the empirical results of this analysis are still relevant and pertinent.

### 4.3. Methodological discussion

This joint spatial marked point process analyses the distribution of individual-level data on dengue cases, adjusting for neighboring effects via spatial structured effects, and accounting for area- and individual-level covariates simultaneously. Given the lack of independence between a dengue case and a severe case (i.e., a severe case needs to be first a case), the advantage of using a joint model to assess the spatial distribution of severe cases relies on several aspects i) the opportunity to use individual location data for overall and severe cases to assess their distribution, ii) the shared common component between the relative risk of cases and the probability of severe dengue that allow us to learn about the underlying spatial process, iii) the opportunity to account for the uncertainty associated with the number of overall dengue cases in the surveillance-based data when modelling the severe cases, and iv) the opportunity of identifying the presence of clustering of severe cases that will otherwise not be identified with separated models for dengue and severe dengue. This approach assumes that there is a spatial trend in the data that cannot be explained by the measured covariates and that such trend is a random field (22, 23, 32).

Although there was a small number of severe dengue cases across the city and within each neighborhood, the joint model assumed the spatial distribution of severe cases proportional to the spatial distribution of overall cases (see equations (2) and (4)) and allowed the identification and confirmation that the spatial patterns of distribution for severe cases showed more dispersion across the city than the distribution of overall cases.

Typically, point process models are fitted using a regular spatial grid which approximates the latent field and the spatial pattern (20, 32). Also, should the data at hand allow it, space-time kernel density estimation (STKDE) could be used for spatiotemporal disease transmission models, which could be computationally intensive and are not necessarily comparable with the methods proposed here (33-36, 41). For ease of applicability among the public health community, data availability, and to avoid issues associated with the interpolation of population offsets, we followed the approach proposed by Pinto Jr. et al.,(36) and used the actual neighborhood map and population information as the spatial grid. This approach facilitated the fitting by providing the real neighboring boundaries and used the actual information of the population, area, and density to improve accuracy. The use of this dataset favors the use and application of research results in the context of surveillance and disease control by decision makers and other stakeholders. We followed Illian et al. (2013)(32) and Pinto Jr. et al., (2015)(36) and approximate the likelihood in the point process model through the neighborhoods of Medellín. Another contribution of our manuscript is to show that the model proposed by Illian et al. (2013), which was developed for an application in ecology, can be considered in the spatial analysis of infectious diseases when data allows. The sensitivity analysis showed some opportunity to gain precision when modeling each case within neighborhoods but further research to identify the spatial dependencies is required. Further development is required to identify alternatives for modelling point process using individual location in infectious diseases.

## Conclusion

These findings provide epidemiological and geographical information of high-risk areas of overall and severe dengue presence in Medellin, Colombia. Age, insurance scheme, and socioeconomic status are key sociodemographic and spatial factors associated with the presence of dengue in the city, but severity was low and without evidence of spatial clustering. The use of joint marked point process models improves the evidence obtained from surveillance data by favoring the assessment of common underlying spatial process, accounting for the uncertainty of overall reported dengue cases, and by favoring its analysis using the observed individual-cases characteristics when data is available. This application contributes to the production of public health information for decision makers to address specific disease control strategies, and to help the preparedness of health services for upcoming outbreaks at the local level.

## Data Availability

Data accessibility: Case-specific data, which is routinely collected using the national surveillance system of Colombia (SIVIGILA; http://portalsivigila.ins.gov.co/sivigila/index.php) was obtained directly form the Local Surveillance office (Secretaria de Salud Municipal de Medellin); Socioeconomic information at neighborhood level was obtained from the website of the municipality (https://www.medellin.gov.co) and an open data source for socioeconomic information (https://www.datos.gov.co/).

## Abbreviations

Cr.Int: Credible Interval
SRR: Standardized Rate Ratio
IQR: Interquartile Range
OR: Odds Ratio
RR: Relative Risk
DIC: Deviation Information Criterion
INLA: Integrated Nested Laplace Approximation

## Data accessibility

Case-specific data, which is routinely collected using the national surveillance system of Colombia (SIVIGILA; http://portalsivigila.ins.gov.co/sivigila/index.php) was obtained directly form the Local Surveillance office (Secretaria de Salud Municipal de Medellin); Socioeconomic information at neighborhood level was obtained from the website of the municipality (https://www.medellin.gov.co) and an open data source for socioeconomic information (https://www.datos.gov.co/).

## Conflict of Interest

The authors declare no conflict of interest

## Funding

This research did not receive any specific grant from funding agencies in the public, commercial, or not-for-profit sectors.

## Acknowledgements

We would like to acknowledge the collaboration of the Secretaria de Salud Municipal de Medellin for their collaboration on accessing the datasets.

## Notes

### Competing Interest Statement

The authors have declared no competing interest.

## REFERENCES

1. Instituto Nacional de Salud (INS). Informe Epidemiologico de Evento Dengue. In: Vigilancia y Análisis del Riesgo en Salud Pública., editor. Bogota, Colombia: Instituto Nacional de Salud; 2016.

2. Villar LA, Rojas DP, Besada-Lombana S, Sarti E. Epidemiological trends of dengue disease in Colombia (2000–2011): a systematic review. PLoS Negl Trop Dis. 2015;9.

3. Pan American Health Organization / World Health Organization. Epidemiological Update: Dengue. Report. Washington, D.C.: PAHO/WHO; 2019 November 11, 2019.

4. World Health Organization (WHO). A Toolkit for National Dengue Burden Estimation. In: World Health Organization (WHO), editor. Geneva2018.

5. Pan American Health Organization (PAHO). Tool for the diagnosis and care of patients with suspected arboviral diseases. 1st Edition ed. Washington: PAHO; 2017 March 2017. 102 p.

6. Mohanty B, Sunder A, Pathak S. Clinicolaboratory profile of expanded dengue syndrome Our experience in a teaching hospital. J Family Med Prim Care. 2019;8(3):1022–7.

7. Waggoner JJ, Gresh L, Mohamed-Hadley A, Balmaseda A, Soda KJ, Abeynayake J, et al. Characterization of Dengue Virus Infections Among Febrile Children Clinically Diagnosed With a Non-Dengue Illness, Managua, Nicaragua. J Infect Dis. 2017;215(12):1816–23.

8. Halstead SB. Antibodies Determine Virulence in Dengue. Annals of the New York Academy of Sciences. 2009;1171:E48–E56.

9. Wearing HJ, Rohani P. Ecological and immunological determinants of dengue epidemics. Proceedings of the National Academy of Sciences. 2006;103(31):11802–7.

10. World Health Organization (WHO). Global strategy for dengue prevention and control 2012-2020. Geneva: WHO. World Health Organization WHO/HTM/NTD; 2012 August 2012. Contract No.: WHO/HTM/NTD/VEM/2012.5.

11. Carabali M, Hernandez L, Arauz M, Villar L, Ridde V. Why are people with dengue dying? A scoping review of determinants for dengue mortality. BMC Infectious Diseases. 2015;15(1):301.

12. Rico-Mendoza A, Alexandra P-R, Chang A, Encinales L, Lynch R. Co-circulation of dengue, chikungunya, and Zika viruses in Colombia from 2008 to 2018. Revista panamericana de salud publica = Pan American journal of public health. 2019;43:e49–e.

13. Martínez-Bello DA, López-Quílez A, Prieto AT. Joint Estimation of Relative Risk for Dengue and Zika Infections, Colombia, 2015-2016. Emerging infectious diseases. 2019;25(6):1118–26.

14. Martínez-Bello DA, López-Quílez A, Torres Prieto A. Relative risk estimation of dengue disease at small spatial scale. International Journal of Health Geographics. 2017;16(1):31.

15. Vanlerberghe V, Gómez-Dantés H, Vazquez-Prokopec G, Alexander N, Manrique-Saide P, Coelho G, et al. Changing paradigms in Aedes control: considering the spatial heterogeneity of dengue transmission. Rev Panam Salud Publica. 2017;41(e16):1–6.

16. Vincenti-Gonzalez MF, Grillet M-E, Velasco-Salas ZI, Lizarazo EF, Amarista MA, Sierra GM, et al. Spatial Analysis of Dengue Seroprevalence and Modeling of Transmission Risk Factors in a Dengue Hyperendemic City of Venezuela. PLOS Neglected Tropical Diseases. 2017;11(1):e0005317.

17. Fritzell C, Rousset D, Adde A, Kazanji M, Van Kerkhove MD, Flamand C. Current challenges and implications for dengue, chikungunya and Zika seroprevalence studies worldwide: A scoping review. PLOS Neglected Tropical Diseases. 2018;12(7):e0006533.

18. Piedrahita LD, Agudelo Salas IY, Marin K, Trujillo AI, Osorio JE, Arboleda-Sanchez SO, et al. Risk Factors Associated with Dengue Transmission and Spatial Distribution of High Seroprevalence in Schoolchildren from the Urban Area of Medellin, Colombia. Canadian Journal of Infectious Diseases and Medical Microbiology. 2018;2018:11.

19. Louis VR, Phalkey R, Horstick O, Ratanawong P, Wilder-Smith A, Tozan Y, et al. Modeling tools for dengue risk mapping - a systematic review. International Journal of Health Geographics. 2014;13(1):1–15.

20. Blangiardo M, Cameletti M. Spatial modeling. Spatial and Spatio-temporal Bayesian Models with R-INLA: John Wiley & Sons, Ltd; 2015. p. 173–234.

21. Congdon P. Models for spatial outcomes and geographical association. Applied Bayesian Modelling: John Wiley & Sons, Ltd; 2014. p. 312–63.

22. Diggle PJ, Moraga P, Rowlingson B, Taylor BM. Spatial and Spatio-Temporal Log-Gaussian Cox Processes: Extending the Geostatistical Paradigm. Statistical Science. 2013;28(4):542–63.

23. Illian JB, Sørbye SH, Rue H. A toolbox for fitting complex spatial point process models using integrated nested Laplace approximation (INLA). The Annals of Applied Statistics. 2012:1499–530.

24. Imai N, Dorigatti I, Cauchemez S, Ferguson NM. Estimating Dengue Transmission Intensity from Sero-Prevalence Surveys in Multiple Countries. PLoS Negl Trop Dis. 2015;9(4):e0003719.

25. Vong S, Goyet S, Ly S, Ngan C, Huy R, Duong V, et al. Under-recognition and reporting of dengue in Cambodia: a capture–recapture analysis of the National Dengue Surveillance System. Epidemiol Infect. 2011;140(3):491–9.

26. Estimaciones de población 1985 - 2005 y proyecciones de población 2005 - 2020 total municipal por área [Internet]. DANE. 2015 [cited September 2018]. Available from: https://www.dane.gov.co/index.php/estadisticas-por-tema/demografia-y-poblacion/proyecciones-de-poblacion.

27. Protocolo de Vigilancia en Salud Pública, Dengue (Surveillance Protocol in Public Health, Dengue)., FOR-R02.0000-059 V02. Sect. V02 (2014).

28. Carabali M, Lim JK, Palencia DC, Lozano-Parra A, Gelvez RM, Lee KS, et al. Burden of dengue among febrile patients at the time of chikungunya introduction in Piedecuesta, Colombia. Tropical Medicine & International Health. 2018;23(0):1231–41.

29. Ley 1438: Por medio de la cual se reforma el Sistema General de Seguridad Social en Salud y se dictan otras disposiciones., (2011.).

30. Departamento Administrativo Nacional de Estadistica (DANE). Metodologia de la Estratificacion Socioeconomica Urbana para Servicios Publicos Domicioliarios. In: Direccion de Geoestadistica, editor. Grupo de Estratificacion ed. Santa Fe de Bogota, Colombia.: DANE; 2015. p. 96.

31. Simpson D, Illian JB, Lindgren F, Sorbye S, Rue H. Going off grid: Computationally efficient inference for log-Gaussian Cox processes. arXiv. 2017:1–26.

32. Illian JB, Martino S, Sorbye S, Gallego-Fernandez JB, Zunzunegui M, Esquivias MP, et al. Fitting complex ecological point process models with Integrated Nested Laplace Approximation. Methods in Ecology and Evolution. 2013;4:305–15.

33. Kang J-Y, Aldstadt J. The Influence of Spatial Configuration of Residential Area and Vector Populations on Dengue Incidence Patterns in an Individual-Level Transmission Model. International Journal of Environmental Research and Public Health. 2017;14(7):792.

34. Hohl A, Delmelle E, Tang W, Casas I. Accelerating the discovery of space-time patterns of infectious diseases using parallel computing. Spatial and Spatio-temporal Epidemiology. 2016;19:10–20.

35. Delmelle E, Dony C, Casas I, Jia M, Tang W. Visualizing the impact of space-time uncertainties on dengue fever patterns. International Journal of Geographical Information Science. 2014;28(5):1107–27.

36. Pinto Junior JA, Gamerman D, Paez MS, Alves Regina HF. Point pattern analysis with spatially varying covariate effects, applied to the study of cerebrovascular deaths. Statistics in Medicine. 2015;34:1214–26.

37. Waller LA, Gotway CA. Analyzing Public Health Data.. In: Shewhart WA, Wilks SS, editors. Applied Spatial Statistics for Public Health Data. 368: John Wiley & Sons; 2004. p. 7–37.

38. Besag J, York J, Mollié A. Bayesian image restoration, with two applications in spatial statistics. Annals of the Institute of Statistical Mathematics. 1991;43(1):1–20.

39. Besag J. Spatial Interaction and the Statistical Analysis of Lattice Systems. Journal of the Royal Statistical Society: Series B (Methodological). 1974;36(2):192–225.

40. Riebler A, Sørbye SH, Simpson D, Rue H. An intuitive Bayesian spatial model for disease mapping that accounts for scaling. Statistical Methods in Medical Research. 2016;25(4):1145–65.

41. Baddeley A, Rubak E, Turner R. Spatial point patterns: methodology and applications with R: CRC Press; 2015.

42. Rue H, Riebler A, Sørbye SH, Illian JB, Simpson DP, Lindgren FK. Bayesian computing with INLA: a review. Annual Review of Statistics and Its Application. 2017;4:395–421.

43. R Core Team. R Foundation for Statistical Computing. Desktop 1.0.143. R version 3.6.0 “Planting of a Tree” ed. Vienna, Austria: R Foundation for Statistical Computing; 2019.

44. Benchimol EI, Smeeth L, Guttmann A, Harron K, Moher D, Petersen I, et al. The REporting of studies Conducted using Observational Routinely-collected health Data (RECORD) Statement. PLOS Medicine. 2015;12(10):e1001885.

45. Simpson D, Rue H, Riebler A, Martins TG, Sørbye SH. Penalising Model Component Complexity: A Principled, Practical Approach to Constructing Priors. Statistical Science. 2017;32(1):1–28,.

46. Restrepo BN, Beatty ME, Goez Y, Ramirez RE, Letson GW, Diaz FJ, et al. Frequency and Clinical Manifestations of Dengue in Urban Medellin, Colombia. Journal of Tropical Medicine. 2014;2014:8.

47. Carabali M, Lim JK, Velez DC, Trujillo A, Egurrola J, Lee KS, et al. Dengue virus serological prevalence and seroconversion rates in children and adults in Medellin, Colombia: implications for vaccine introduction. International Journal of Infectious Diseases. 2017;58:27–36.

48. Boyer S, Foray C, Dehecq J-S. Spatial and temporal heterogeneities of Aedes albopictus density in La Reunion Island: rise and weakness of entomological indices. PLoS One. 2014;9.

49. Chiaravalloti-Neto F, Pereira M, Fávaro EA, Dibo MR, Mondini A, Rodrigues-Junior AL, et al. Assessment of the relationship between entomologic indicators of Aedes aegypti and the epidemic occurrence of dengue virus 3 in a susceptible population, São José do Rio Preto, São Paulo, Brazil. Acta Tropica. 2015;142:167–77.

50. Ocampo CB, Mina NJ, Carabalí M, Alexander N, Osorio L. Reduction in dengue cases observed during mass control of Aedes (Stegomyia) in street catch basins in an endemic urban area in Colombia. Acta Tropica. 2014;132(0):15–22.

51. Ardila Pinto F, Martínez S, Fuentes M, Borrero E. Análisis de las demoras en salud en personas que enfermaron de gravedad o fallecieron por dengue en cinco ciudades de Colombia. Physis: Revista de Saúde Coletiva. 2015;25:571–92.

52. Carabalí JM, Hendrickx D. Dengue and health care access: the role of social determinants of health in dengue surveillance in Colombia. Glob Health Promot. 2012;19.

53. Arauz MJ, Ridde V, Hernández LM, Charris Y, Carabali M, Villar LÁ. Developing a Social Autopsy Tool for Dengue Mortality: A Pilot Study. PLoS ONE. 2015;10(2):e0117455.

54. Brian JR, Hodges JS, Vesna Z. Effects of Residual Smoothing on the Posterior of the Fixed Effects in Disease-Mapping Models. Biometrics. 2006;62(4):1197–206.

55. Wichmann Ole, Yoon IK, Vong S, Limkittikul K, Gibbons RV, Mammen PM, et al. Dengue in Thailand and Cambodia: An Assessment of the Degree of Underrecognized Disease Burden Based on Reported Cases. PLoS Negl Trop Dis. 2011;5(3):e996.

