## Supplementary Material for "A joint spatial marked point process model for dengue and severe dengue in Medellin, Colombia"

Here we present some additional descriptive information about the distribution of dengue cases, additional details about spatial point process models, detailed results from complementary analysis and results of additional sensitivity analysis.

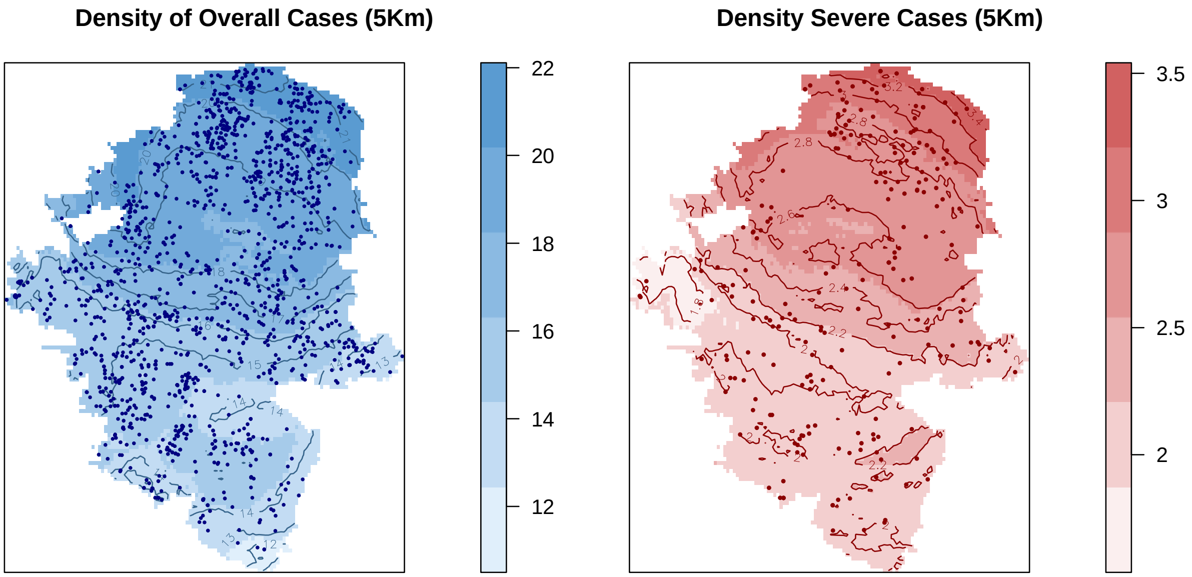


**Figure S1.** Distribution of dengue cases notified in Medellin in 2013. Kernel density of overall and severe dengue cases using a 5 Km bandwidth. Solid circles indicate a case and the darker the color (blue for overall dengue cases and red for severity) indicates higher density of cases per 5 $Km^2$.

### 1. Spatial point process model:

A log-Gaussian Cox point process model assesses the distribution of the individual location of the outcome points (dengue cases) in a spatial structure, and it is used in order to consider both, observed and unobserved variation in the assessment of such distribution[1]. It estimates the spatial distribution of the individual cases as a function of a continuous latent Gaussian random field, assuming conditional independence of the points presented on the field. This indicates that conditional on the latent field, the distribution of the point pattern (dengue cases), follows a Poisson process[1-3]. This analysis uses individual level information and allows covariates at the spatial level to vary according to the random field. Thus, providing information about the presence and degree of clustering within the spatial structure, while considering simultaneously the spatial autocorrelation between and within spatial units[4].

A Marked log-Gaussian Cox point process model uses an individual characteristic of the point, the 'mark', to assess the individual distribution of an event given such specific characteristic. In our case, each point represents an individual case, and the "mark" is the presence/absence of severity for each dengue case. Thus, the information about the "mark" is used to estimate two aspects simultaneously: 1) the overall distribution of the point pattern (i.e., all points/cases) and 2) the distribution of the point pattern given the mark (i.e., severe cases). The model could also be considered a "labeling" of the Poisson process, where the 'marks' work also as a response variable[2]. This approach is intended to identify whether there is an underlying mechanism leading to a differential distribution of severe cases, with the added advantage of modelling simultaneously an individual feature of the point (the severity) and the spatial distribution, while accounting for its dependence[2,3].

To conduct a point process analysis, it is necessary to consider each case-location as $x_i: i = 1, \ldots n$, where, $x$ indicates the location and $i$ indicates the dengue case identifier that in theory could have occurred in any location inside a given spatial region $A \subset \Re^2$ [1]. Likewise, it is important to accept two main structural assumptions: i) that the function of the Cox-Process is a stochastic non-negative process $\Lambda = \{\Lambda(x): x \in \Re^2\}$, and ii) that conditional on the realization $\Lambda(x) = \lambda(x): x \in \Re^2$, the point process is an inhomogeneous Poisson process with intensity $\lambda(x)$. Also, it is necessary to consider the distribution of dengue cases as a phenomenon $S(x)$ that is incompletely observed and spatially continuous, given that $S = \{S(x) : x \in \Re^2\}$ is a Gaussian stochastic process and that $S$ determines $\lambda(x)$, which is the intensity of the distribution; $\lambda(x) = \exp\{S(x)\}$[1,3]. To analyze this intensity, it is necessary to approximate the spatially continuous random field to a constructed grid [1,3,5]. Then, considering $\{y_i\}$ the observed number of points in the neighborhood, we assume that the number of points (cases) in a grid-cell/neighborhood $i$ follows a Poisson distribution conditional on a first latent field, $\eta_i^{(1)}$:

$$y_i|\eta_i^{(1)} \sim Po\left(E_i\ \eta_i^{(1)}\right), \text{ Equation (S.1)}$$

The offset of the pattern $E_i$ is specified as the expected count of cases in each neighborhood and obtained via indirect standardization[6]. To model the marked point process, we add to equation 1, the analysis of the marks (severe or not severe). For that we let $m_i$ be the number of patients with severe dengue in each spatial unit $i$ . Then, conditional on the value of a second latent field $\eta_i^{(2)}$ in the same neighborhood, $m_i$ follows a binomial distribution:

$$m_i|\eta_i^{(2)} \sim Binomial\left(y_i,\ \eta_i^{(2)}\right), \text{ Equation (S.2)}$$

where $\eta_i^{(2)}$is the probability of being a severe case in a given neighborhood $i$ while $y_i$ is the total number of cases of dengue in neighborhood $i$. This, constituting a matrix outcome of two links (i.e., Poisson for overall point patterns, and Binomial for severity), each one on a separate latent field $\eta_i^{(j)}$, that are jointly analyzed in relation to a vector of sociodemographic covariates[2]. We constructed the final model for each latent field $\eta_i^{(1)}$ and $\eta_i^{(2)}$, including empirical covariates for neighborhood level and observed case characteristics as fixed effects, and spatial structures as random effects as follows:

$$log\left(\ \eta_i^{(1)}\right) = \beta_0^{(1)} + \beta_1^{(1)} IB(s_i) + \beta_2^{(1)} UNDER20(s_i) + \beta_3^{(1)} P.FEMALE(s_i) + \\ \beta_4^{(1)} P.INSURANCE(s_i)\ + \beta_5^{(1)} SES(s_i) + f_s^{j}(s_i) + u(s_i), \text{ Equation (S.3)}$$

$$logit\left(\eta_i^{(2)}\right) = \beta_0^{(2)} + \beta_1^{(2)} AGE1(s_i) + \beta_2^{(2)} AGE2(s_i) + \beta_3^{(2)} SEX(s_i) + \beta_4^{(2)} INSURANCE(s_i) + \\ \beta_5^{(2)} DISTANCEKm(s_i)\ + g_s^{j}(s_i) + v(s_i), \text{ Equation (S.4)}$$

$\beta_0^{(1)}$ and$\beta_0^{(2)}$ are the pattern and marks intercepts. $\beta_1^{(1)}$ … $\beta_4^{(1)}$ are the coefficients associated to the empirical covariates for the distribution of cases at the neighborhood level; and $\beta_1^{(2)}$ … $\beta_4^{(2)}$ are the coefficients associated to the empirical covariates for severity at the individual level, as described in the main text. $DISTANCEKm(s_i)$ is the standardized minimum nearest-neighbor Euclidean distance between overall and severe cases, parameterized as a continuous variable. The components $f_s^j(s_i)$ and $g_s^j(s_i)$ are the Gaussian Markov Random Field (GMRF), reflecting separately the spatial autocorrelation in the latent field, working as spatially structured effects for the pattern and the marks, respectively. $u(s_i)$ is the spatially unstructured random effect for the pattern and $v(s_i)$ is the spatially unstructured random effect for the marks. To express the dependence between the pattern and the marks, we used a single (common) random field replacing equation (4) as follows:

$$logit\left(\eta_i^{(2)}\right) = \beta_0^{(2)} + \beta_1^{(2)}AGE1(s_i) + \beta_2^{(2)}AGE2(s_i) + \beta_3^{(2)}SEX(s_i) + \beta_4^{(2)}INSURANCE(s_i) + \beta_5^{(2)}DISTANCEKm(s_i) + \beta_s f_s^i(s_i) + v(s_i), \text{ Equation (S.5)}$$

which makes the spatial effect for the severity proportional to the spatial effect of the pattern of case distribution [2]. The spatially structured effects were captured by assuming the Besag specification and the models including the structured and unstructured effects simultaneously were modeled following the Besag-York-Mollie (BYM) specification[7]; where after adjusting for the fixed effects, the structured component is modeled using an intrinsic conditional autoregressive structure (iCAR) and the unstructured effect is modeled using an independent prior[6,8,9] and the BYM2 specification, which is a reparameterization of the BYM and included a mixing parameter (Phi) with the interpretation as an indicator of spatial dependency, which helps understanding the decomposition between spatially structured and independent random effects[10].

Below we provide the results for single models for the pattern (Table S1) and the severity (Table S2a and S2b) and the joint proportional models (Tables S3a and S3b), using each one of the Besag, BYM and BYM2 parameterizations of the spatial structure. Table S4 shows the model without including SES variable. Table S5 shows the DIC for different model specification for the joint proportional models using the Besag parameterizations of the spatial structure. Figure S2 shows the posterior density of fixed and random effects of the final joint model using a single spatial component and Figure S3 shows the maps of the spatial distribution the final model under different specifications.

**Table. S1.** Posterior mean of the Standardized Rate Ratio (SRR)mand 95% credible intervals for covariates (fixed effects) on the **single-separated model for overall dengue** cases in Medellin, 2013.

| | **Besag (DIC=1214.75)** | | | **BYM (DIC=1214.75)** | | | **BYM2 (DIC=1214.42)** | | |
|---|---|---|---|---|---|---|---|---|---|
| **Patterns' Covariates** | **SRR** | **2.5%** | **97.5%** | **SRR** | **2.5%** | **97.5%** | **SRR** | **2.5%** | **97.5%** |
| (Intercept) | 0.37 | 0.24 | 0.56 | 0.33 | 0.21 | 0.50 | -1.09 | -1.54 | -0.65 |
| Proportion of **female cases** | 1.01 | 1.00 | 1.01 | 1.01 | 1.00 | 1.01 | 0.01 | 0.00 | 0.01 |
| Proportion of **cases <20 years** old | 1.01 | 1.00 | 1.01 | 1.01 | 1.01 | 1.02 | 0.01 | 0.01 | 0.02 |
| Proportion of **contributory scheme** cases | 1.01 | 1.00 | 1.01 | 1.01 | 1.00 | 1.01 | 0.01 | 0.00 | 0.01 |
| **Medium** SES | Ref. | - | - | Ref. | - | Ref. | - | - | |
| **Low** SES | 0.63 | 0.44 | 0.88 | 0.56 | 0.42 | 0.75 | -0.54 | -0.84 | -0.23 |
| **High** SES | 0.64 | 0.42 | 0.98 | 0.68 | 0.46 | 0.98 | -0.48 | -0.89 | -0.08 |
| Medium **Breteau Index** | 1.02 | 0.79 | 1.30 | 0.95 | 0.74 | 1.21 | -0.05 | -0.30 | 0.20 |
| **Hyperparameter** | **Mean** | **2.5%** | **97.5%** | **Mean** | **2.5%** | **97.5%** | **Mean** | **2.5%** | **97.5%** |
| Precision for $f_s^j(s_i)$ | 0.749 | 0.568 | 0.969 | | | | 1.508 | 1.14 | 1.97 |
| Precision for $u_{(s_i)}$ | | | | 1.53 | 1.17 | 1.97 | | | |
| Precision for $f_s^j(s_i) + u_{(s_i)}$ | | | | 2109.84 | 121.69 | 7867.26 | | | |
| Phi for $u_{(s_i)}$ | | | | | | | 0.173 | 0.02 | 0.47 |

Proportion of woman, proportion of cases under 20 years old, and proportion of contributory scheme indicate 10% increase. Breteau Index reference group = Low. SES Level: Comparing cases in the Medium SES (Reference group) to Cases in the Low and High SES levels.

**Table. S2a.** Posterior mean of the Odds Ratio and 95% credible intervals for covariates (fixed effects) on the **single-separated model** for **severe dengue** cases in Medellin, 2013. Proportional model without Neighborhood-SES covariate.

| | **Besag (DIC =367.93)** | | | **BYM (DIC=367.81)** | | | **BYM2 (DIC=368.61)** | | |
|---|---|---|---|---|---|---|---|---|---|
| **Severity Covariates** | **OR** | **2.5%** | **97.5%** | **OR** | **2.5%** | **97.5%** | **OR** | **2.5%** | **97.5%** |
| (Intercept) | 0.17 | 0.04 | 0.88 | 0.17 | 0.04 | 0.89 | 0.17 | 0.04 | 0.92 |
| Proportion of f**emale** cases | 1.00 | 0.99 | 1.00 | 1.00 | 0.99 | 1.00 | 1.0 | 0.99 | 1.0 |
| Proportion of **cases <20 years** old | 1.00 | 0.99 | 1.02 | 1.00 | 0.99 | 1.02 | 1.0 | 0.99 | 1.02 |
| Proportion of **cases >20 years** old | 1.01 | 0.99 | 1.02 | 1.01 | 0.99 | 1.02 | 1.01 | 0.99 | 1.02 |
| Proportion of **contributory scheme** cases | 1.00 | 1.00 | 1.00 | 1.00 | 1.00 | 1.00 | 1.0 | 1.0 | 1.0 |
| **Median distance** to Severe cases (Km) | 1.00 | 1.00 | 1.00 | 1.00 | 1.00 | 1.00 | 1.0 | 1.0 | 1.0 |
| **Hyperparameters** | **Mean** | **2.5%** | **97.5%** | **Mean** | **2.5%** | **97.5%** | **Mean** | **2.5%** | **97.5%** |
| Precision for $f_s^j(s_i)$ | 18782.56 | 524.23 | 64824.22 | | | | 508.52 | 12.23 | 3281.20 |
| Precision for $u_{(s_i)}$ | | | | 2016.66 | 151.32 | 7424.19 | 0.331 | 0.007 | 0.93 |
| Precision for $f_s^j(s_i) + u_{(s_i)}$ | | | | 2162.17 | 189.03 | 7892.38 | | | |
| Phi for $u_{(s_i)}$ | | | | | | | 0.331 | 0.007 | 0.93 |

Proportion of woman, proportion of cases under (or over) 20 years old, and proportion of contributory scheme indicate 10% increase.
SES Level: Comparing cases in the Medium SES (Reference group) to Cases in the Low and High SES levels.

**Table S2b.** Posterior mean of the Odds Ratio and 95% credible intervals for covariates (fixed effects) on the **single-separated model for severe dengue** cases in Medellin, 2013. Proportional model with Neighborhood-SES covariate.

| | **Besag (DIC =** 371.35**)** | | | **BYM (DIC=** 371.17) | | | **BYM2 (DIC=**371.94) | | |
|---|---|---|---|---|---|---|---|---|---|
| **Severity Covariates** | **OR** | **2.5%** | **97.5%** | **OR** | **2.5%** | **97.5%** | **OR** | **2.5%** | **97.5%** |
| Intercept | 0.19 | 0.04 | 1.00 | 0.19 | 0.04 | 1.01 | 0.19 | 0.04 | 1.05 |
| Proportion of **female** cases | 1.00 | 0.99 | 1.00 | 1.00 | 0.99 | 1.00 | 1.00 | 0.99 | 1.00 |
| Proportion of **cases <20 years** old | 1.00 | 0.99 | 1.02 | 1.00 | 0.99 | 1.02 | 1.00 | 0.99 | 1.02 |
| Proportion of **cases >20 years** old | 1.01 | 0.99 | 1.02 | 1.01 | 0.99 | 1.02 | 1.01 | 0.99 | 1.02 |
| Proportion of **contributory scheme** cases | 1.00 | 0.99 | 1.00 | 1.00 | 0.99 | 1.00 | 1.00 | 0.99 | 1.00 |
| **Medium** SES | Ref. | - | - | Ref. | - | - | Ref. | - | - |
| **Low** SES | 0.94 | 0.68 | 1.28 | 0.94 | 0.68 | 1.28 | 0.93 | 0.67 | 1.29 |
| **High** SES | 1.14 | 0.70 | 1.83 | 1.14 | 0.70 | 1.83 | 1.13 | 0.68 | 1.85 |
| **Median Distance** to Severe cases (Km) | 1.00 | 1.00 | 1.00 | 1.00 | 1.00 | 1.00 | 1.00 | 1.00 | 1.00 |
| **Hyperparameter** | **Mean** | **2.5%** | **97.5%** | **Mean** | **2.5%** | **97.5%** | **Mean** | **2.5%** | **97.5%** |
| Precision for $f_s^j(s_i)$ | 18759.39 | 510.83 | 64801.49 | | | | | | |
| Precision for $u_{(s_i)}$ | | | | 2057.30 | 158.44 | 7611.9 | | | |
| Precision for $f_s^j(s_i) + u_{(s_i)}$ | | | | 2197.06 | 194.96 | 8051.27 | 410.96 | 11.04 | 2662.18 |
| Phi for $u_{(s_i)}$ | | | | | | | 0.337 | 0.007 | 0.933 |

Proportion of woman, proportion of cases under (or over) 20 years old, and proportion of contributory scheme indicate 10% increase.
SES Level: Comparing cases in the Medium SES (Reference group) to Cases in the Low and High SES levels.

**Table S3a.** Posterior mean of the Standardized Rate Ratio (SRR), Odds Ratios (OR) and 95% credible intervals of covariates (fixed effects) for **the joint proportional model for overall dengue distribution and severity**.

| | Besag (DIC=1578.22) | | | BYM (DIC=1572.58) | | | BYM2 (DIC=1572.5) | | |
|---|---|---|---|---|---|---|---|---|---|
| **Pattern's Covariates** | **SRR** | **2.5%** | **97.5%** | **SRR** | **2.5%** | **97.5%** | **SRR** | **2.5%** | **97.5%** |
| Proportion of **female cases** | 1.07 | 1.03 | 1.12 | 1.08 | 1.04 | 1.13 | 1.08 | 1.03 | 1.13 |
| Proportion of **cases <20 years** old | 1.10 | 1.04 | 1.16 | 1.12 | 1.06 | 1.18 | 1.12 | 1.06 | 1.18 |
| Proportion of **contributory scheme** cases | 1.07 | 1.02 | 1.12 | 1.09 | 1.04 | 1.14 | 1.08 | 1.03 | 1.13 |
| **Medium** SES | Ref. | - | - | Ref. | - | - | Ref. | - | - |
| **Low** SES | 0.74 | 0.56 | 0.98 | 0.67 | 0.5 | 0.89 | 0.68 | 0.51 | 0.91 |
| **High** SES | 0.69 | 0.48 | 1.01 | 0.66 | 0.45 | 0.96 | 0.64 | 0.44 | 0.93 |
| Medium **Breteau Index** | 1.06 | 0.83 | 1.36 | 1.01 | 0.78 | 1.29 | 1.02 | 0.79 | 1.31 |
| **Severity Covariates** | **OR** | **2.5%** | **97.5%** | **OR** | **2.5%** | **97.5%** | **OR** | **2.5%** | **97.5%** |
| Proportion of **female** cases | 0.97 | 0.93 | 1.01 | 0.97 | 0.93 | 1.01 | 0.97 | 0.93 | 1.01 |
| Proportion of **cases <20 years** old | 1.04 | 0.88 | 1.2 | 1.04 | 0.88 | 1.2 | 1.04 | 0.88 | 1.2 |
| Proportion of **cases >20 years** old | 1.06 | 0.9 | 1.23 | 1.06 | 0.9 | 1.23 | 1.06 | 0.9 | 1.23 |
| Proportion of **contributory scheme** cases | 0.99 | 0.95 | 1.03 | 0.99 | 0.95 | 1.03 | 0.99 | 0.95 | 1.03 |
| **Median distance** to Severe cases (Km) | 0.99 | 0.98 | 1.00 | 0.99 | 0.990 | 1.000 | 0.99 | 0.99 | 1.00 |
| **Hyperparameter** | **Mean** | **2.5%** | **97.5%** | **Mean** | **2.5%** | **97.5%** | **Mean** | **2.5%** | **97.5%** |
| Precision for $f_s^j(s_i)$ | 0.331 | 0.25 | 0.428 | | | | 1.452 | 1.16 | 1.844 |
| $\exp$ ($\beta$ Coefficient for Severity for $g_s^j(s_{ij})$) | 0.78 | 0.65 | 0.93 | 0.79 | 0.66 | 0.95 | 0.79 | 0.66 | 0.93 |
| Precision for $u_{(s_i)}$ | | | | 1.55 | 1.173 | 2.007 | | | |
| Precision for $f_s^j(s_i) + u_{(s_i)}$ | | | | NC | NC | NC | | | |
| Phi for $u_{(s_i)}$ | | | | | | | 0.223 | 0.032 | 0.568 |

*Proportion of woman, proportion of cases under 20 years old, and proportion of contributory scheme indicate 10% increase. Breteau Index reference group = Low; Age group reference= <15 years of age; Sex reference= Male; Health Insurance reference is Subsidized scheme. SES Level: Comparing cases in the Medium SES (Reference group) to Cases in the Low and High SES levels.NC=No convergence.*

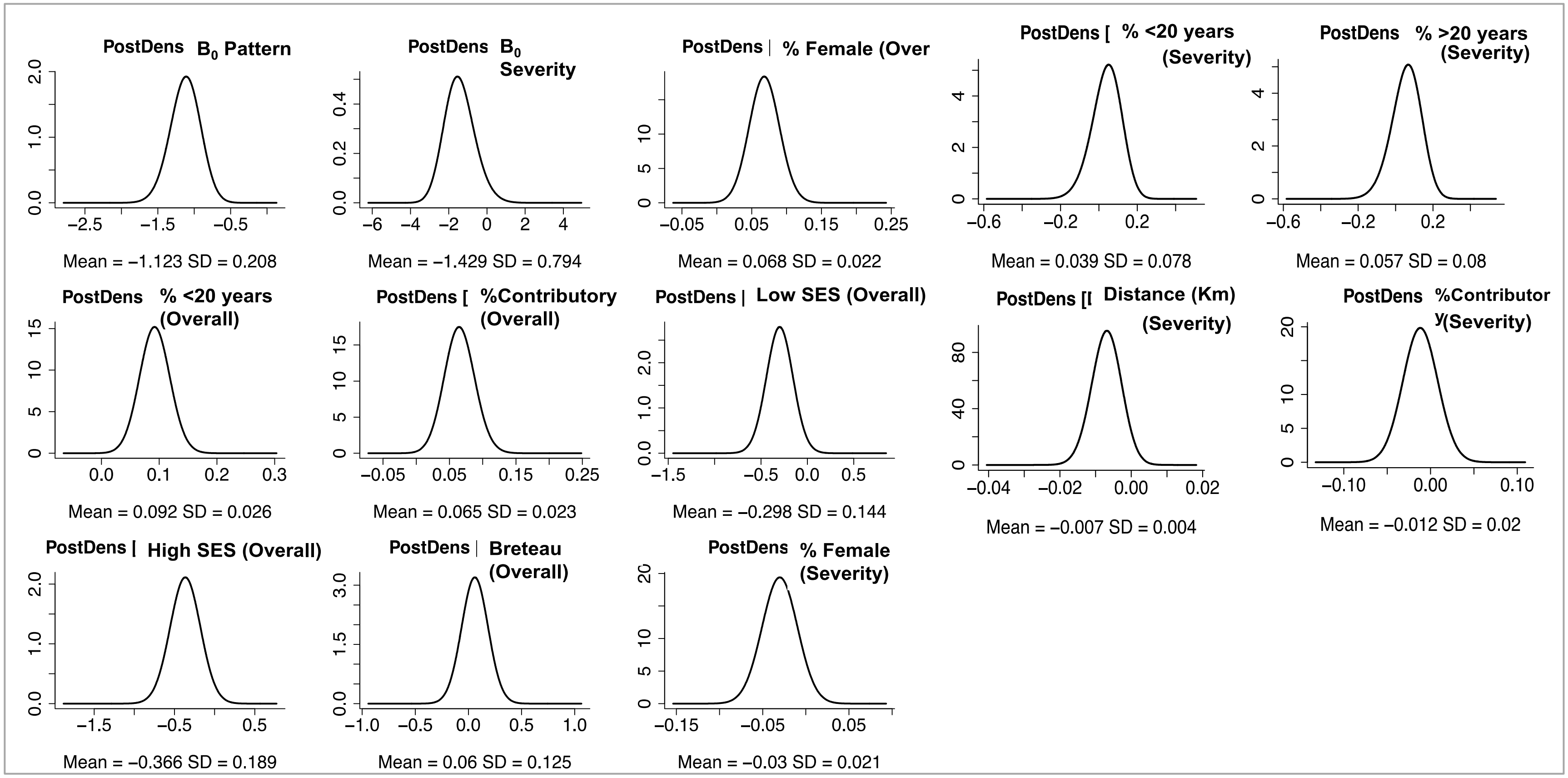


Figure S2. Posterior density of fixed and random effects of the final joint model using a single spatial component

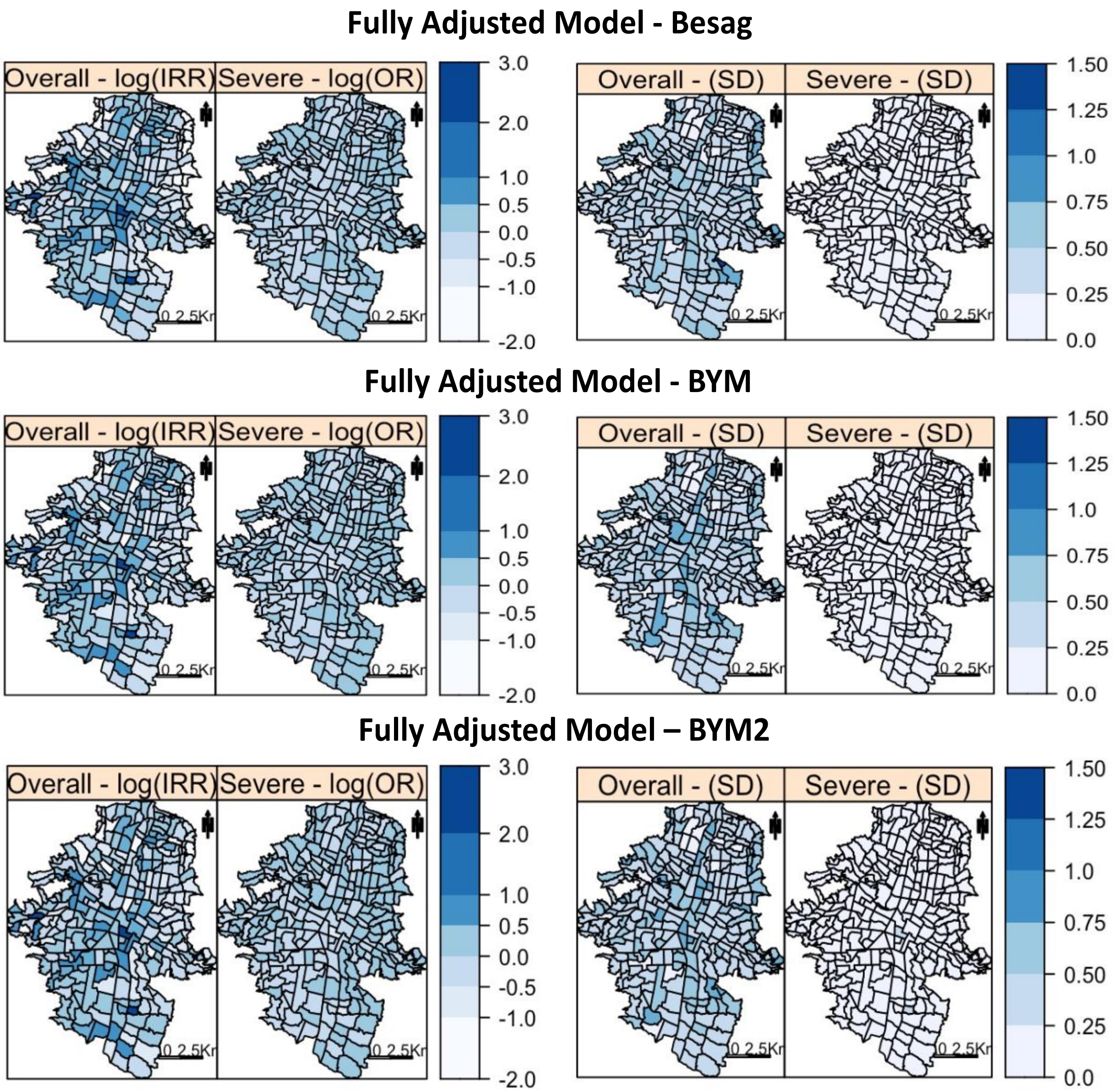


Figure S3. Estimated common spatial trend for overall dengue and severe dengue cases in Medellin, 2013. Neighborhood specific residual (Random Effects) Standardized Rate Ratio (nSRR) and Standard Deviation (SD). Neighborhood specific residual (Random Effects) Odds Ratio (nOR) and Standard Deviation (SD).

**Table S3b.** Posterior mean of the Standardized Rate Ratio (SRR), Odds Ratios (OR) and 95% credible intervals of covariates (fixed effects) for **the joint proportional model for overall dengue distribution and severity, including SES for severity assessment**.

| | Besag (DIC=1575.79) | | | BYM (DIC =1575.79) | | | BYM2 (DIC =1581.36) | | |
|---|---|---|---|---|---|---|---|---|---|
| **Patterns' Covariats** | **SRR** | **2.5%** | **97.5%** | **SRR** | **2.5%** | **97.5%** | **SRR** | **2.5%** | **97.5%** |
| Proportion of **female cases** | 1.07 | 1.03 | 1.12 | 1.08 | 1.04 | 1.13 | 1.08 | 1.03 | 1.13 |
| Proportion of **cases <20 years** old | 1.1 | 1.04 | 1.16 | 1.12 | 1.06 | 1.18 | 1.12 | 1.06 | 1.18 |
| Proportion of **contributory scheme** cases | 1.07 | 1.02 | 1.12 | 1.09 | 1.04 | 1.14 | 1.08 | 1.03 | 1.13 |
| **Medium** SES | Ref. | - | - | Ref. | - | - | Ref. | - | - |
| **Low** SES | 0.75 | 0.56 | 0.99 | 0.66 | 0.5 | 0.89 | 0.68 | 0.51 | 0.92 |
| **High** SES | 0.68 | 0.47 | 0.99 | 0.65 | 0.44 | 0.95 | 0.63 | 0.43 | 0.92 |
| Medium **Breteau Index** | 1.06 | 0.83 | 1.36 | 1.01 | 0.78 | 1.29 | 1.02 | 0.79 | 1.31 |
| **Severity Covariates** | **OR** | **2.5%** | **97.5%** | **OR** | **2.5%** | **97.5%** | **OR** | **2.5%** | **97.5%** |
| Proportion of **female** cases | 0.97 | 0.93 | 1.01 | 0.97 | 0.93 | 1.01 | 0.97 | 0.93 | 1.01 |
| Proportion of **cases <20 years** old | 1.03 | 0.88 | 1.19 | 1.03 | 0.88 | 1.19 | 1.03 | 0.88 | 1.19 |
| Proportion of **cases >20 years** old | 1.05 | 0.89 | 1.22 | 1.05 | 0.89 | 1.22 | 1.05 | 0.89 | 1.22 |
| Proportion of **contributory scheme** cases | 0.98 | 0.94 | 1.03 | 0.98 | 0.94 | 1.03 | 0.98 | 0.94 | 1.03 |
| **Medium** SES | Ref. | - | - | Ref. | - | - | Ref. | - | - |
| **Low** SES | 0.97 | 0.7 | 1.34 | 1 | 0.72 | 1.38 | 0.99 | 0.71 | 1.37 |
| **High** SES | 1.17 | 0.71 | 1.89 | 1.18 | 0.72 | 1.9 | 1.19 | 0.73 | 1.92 |
| **Median Distance** to Severe cases (Km) | 0.99 | 0.98 | 1 | 0.99 | 0.98 | 1 | 0.99 | 0.98 | 1 |
| **Hyperparameter** | **Mean** | **2.5%** | **97.5%** | **Mean** | **2.5%** | **97.5%** | **Mean** | **2.5%** | **97.5%** |
| Precision for $f_s^j(s_i)$ | 0.331 | 0.25 | 0.429 | | | | 1.43 | 1.1 | 1.849 |
| $\exp$ ($\beta$ Coefficient for Severity for $g_s^j(s_{ij})$) | 0.78 | 0.65 | 0.93 | 0.79 | 0.66 | 0.94 | 0.79 | 0.66 | 0.94 |
| Precision for $u_{(s_i)}$ | | | | 1.549 | 1.173 | 2.006 | | | |
| Precision for $f_s^j(s_i) + u_{(s_i)}$ | | | | NC | 0 | NC | | | |
| Phi for $u_{(s_i)}$ | | | | | | | 0.205 | 0.030 | 0.498 |

*Proportion of woman, proportion of cases under 20 years old, and proportion of contributory scheme indicate 10% increase. Breteau Index reference group = Low; Age group reference= <15 years of age; Sex reference= Male; Health Insurance reference is Subsidized scheme. SES Level: Comparing cases in the Medium SES (Reference group) to Cases in the Low and High SES levels. NC=No convergence.*

**Table S4.** Standardized Rate Ratio (SRR), Odds Ratios (OR) and 95% credible intervals of covariates (fixed effects) for the model **without the SES covariates**, Besag (DIC=1577.97).

| | Estimates and 95% Credible intervals (DIC=1577.97) | | |
|---|---|---|---|
| **Overall Cases (Pattern) Covariates** | **SRR** | **2.5%** | **97.5%** |
| Proportion of **female cases** | 1.07 | 1.03 | 1.12 |
| Proportion of **cases <20 years** old | 1.09 | 1.03 | 1.14 |
| Proportion of **contributory scheme** cases | 1.07 | 1.03 | 1.12 |
| Medium **Breteau Index** | 1.04 | 0.81 | 1.33 |
| **Severity Covariates** | **OR** | **2.5%** | **97.5%** |
| Proportion of **female** cases | 0.97 | 0.93 | 1.01 |
| Proportion of **cases <20 years** old | 1.04 | 0.88 | 1.2 |
| Proportion of **cases >20 years** old | 1.06 | 0.89 | 1.22 |
| Proportion of **contributory scheme** cases | 0.99 | 0.95 | 1.03 |
| **Median Distance** to Severe cases (Km) | 0.99 | 0.98 | 1.00 |
| **Hyperparameter** | mean | **2.5%** | **97.5%** |
| Precision for $f_s^j(s_i)$ | 0.32 | 0.243 | 0.413 |
| **Spatial Effect** $\exp$ ($\beta_s$ Coefficient for Severity for $g_s^j(s_{ij})$) | 0.785 | 0.658 | 0.936 |

*Area Level Covariates: Proportion of Female Cases: indicates every 10% increase in the proportion of female cases reported per neighborhood; Proportion of cases <20 years old: indicates every 10% increase in the proportion of reported cases <20 years old per neighborhood. Proportion of Contributory Scheme Cases: indicates every 10% increase in the proportion of cases with contributory scheme insurance reported per neighborhood; Breteau Index: Comparing the Low Breteau Index level (Reference group) to Medium Breteau Index level.*

**Table S5.** Summary of Deviance Information Criterion (DIC) values and specification for joint models of overall (pattern) and severe (marks) dengue cases using Besag parameterization.

| Model | Model Components | DIC |
|---|---|---|
| Model **without covariates** | $log(\ \eta_i^{(1)}) = \beta_0^{(1)} + f_s^j(s_i) + u(s_i);$ <br> $logit(\eta_i^{(2)}) = \beta_0^{(2)} + \beta_s f_s^i(s_i)$ | 1605.8 |
| Model with **only <20 years of age** for severity. | $log(\ \eta_i^{(1)}) = \beta_0^{(1)} + \beta_1^{(1)} IB(s_i) + \beta_2^{(1)} UNDER20(s_i) + \beta_3^{(1)} P.INSURANCE(s_i) + \beta_4^{(1)} P.FEMALE(s_i) + \ \beta_4^{(1)} SES(s_i) + f_s^j(s_i) + u(s_i);$ <br> $logit(\eta_i^{(2)}) = \beta_0^{(2)} + \beta_1^{(2)} AGE1(s_i) + \beta_2^{(2)} SEX(s_i) + \beta_3^{(2)} INSURANCE(s_i) + \beta_4^{(2)} DISTANCEKm(s_i)\ + \beta_s f_s^i(s_i) + v(s_i),$ | 1576.6 |
| Model with **only >20 years of age** for severity. | $log(\ \eta_i^{(1)}) = \beta_0^{(1)} + \beta_1^{(1)} IB(s_i) + \beta_2^{(1)} UNDER20(s_i) + \beta_3^{(1)} P.INSURANCE(s_i) + \beta_4^{(1)} P.FEMALE(s_i) + \ \beta_4^{(1)} SES(s_i) + f_s^j(s_i) + u(s_i);$ <br> $logit(\eta_i^{(2)}) = \beta_0^{(2)} + \beta_1^{(2)} AGE2) + \beta_2^{(2)} SEX(s_i) + \beta_3^{(2)} INSURANCE(s_i) + \beta_4^{(2)} DISTANCEKm(s_i)\ + \beta_s f_s^i(s_i) + v(s_i),$ | 1576.4 |
| Full model **without any SES covariates.** | $log(\ \eta_i^{(1)}) = \beta_0^{(1)} + \beta_1^{(1)} IB(s_i) + \beta_2^{(1)} UNDER20(s_i) + \beta_3^{(1)} P.INSURANCE(s_i) + \beta_4^{(1)} P.FEMALE(s_i) + f_s^j(s_i) + u(s_i);$ <br> $logit(\eta_i^{(2)}) = \beta_0^{(2)} + \beta_1^{(2)} AGE1(s_i) + \beta_2^{(2)} AGE2(s_i) + \beta_3^{(2)} SEX(s_i) + \beta_4^{(2)} INSURANCE(s_i) + \beta_5^{(2)} DISTANCEKm(s_i)\ + \beta_s f_s^i(s_i) + v(s_i),$ | 1577.9 |
| Full model **without the SES covariate for severity**. | $log(\ \eta_i^{(1)}) = \beta_0^{(1)} + \beta_1^{(1)} IB(s_i) + \beta_2^{(1)} UNDER20(s_i) + \beta_3^{(1)} P.INSURANCE(s_i) + \beta_4^{(1)} P.FEMALE(s_i) + \ \beta_4^{(1)} SES(s_i) + f_s^j(s_i) + u(s_i);$ <br> $logit(\eta_i^{(2)}) = \beta_0^{(2)} + \beta_1^{(2)} AGE1(s_i) + \beta_2^{(2)} AGE2(s_i) + \beta_3^{(2)} SEX(s_i) + \beta_4^{(2)} INSURANCE(s_i) + \beta_5^{(2)} DISTANCEKm(s_i)\ + \beta_s f_s^i(s_i) + v(s_i),$ | 1578.2 |
| Full model **without the SES cov**ariate for severity & **two separate spatial structures.** | $log(\ \eta_i^{(1)}) = \beta_0^{(1)} + \beta_1^{(1)} IB(s_i) + \beta_2^{(1)} UNDER20(s_i) + \beta_3^{(1)} P.FEMALE(s_i) + \beta_4^{(1)} SES(s_i) + f_s^j(s_i) + u(s_i);$ <br> $logit(\eta_i^{(2)}) = \beta_0^{(2)} + \beta_1^{(2)} AGE1(s_i) + \beta_2^{(2)} AGE2(s_i) + \beta_3^{(2)} SEX(s_i) + \beta_4^{(2)} INSURANCE(s_i) + \beta_5^{(2)} DISTANCEKm(s_i)\ + g_s^{(2)}(s_i) + v(s_i)$ | 1590 |
| Full model with the **SES covariate for severity**. | $log(\ \eta_i^{(1)}) = \beta_0^{(1)} + \beta_1^{(1)} IB(s_i) + \beta_2^{(1)} UNDER20(s_i) + \beta_3^{(1)} P.INSURANCE(s_i) + \beta_4^{(1)} P.FEMALE(s_i) + \ \beta_4^{(1)} SES(s_i) + f_s^j(s_i) + u(s_i);$ <br> $logit(\eta_i^{(2)}) = \beta_0^{(2)} + \beta_1^{(2)} AGE1(s_i) + \beta_2^{(2)} AGE2(s_i) + \beta_3^{(2)} SEX(s_i) + \beta_4^{(2)} INSURANCE(s_i) + \beta_5^{(2)} DISTANCEKm(s_i) + \ \beta_6^{(2)} SES(s_i) + \beta_s f_s^i(s_i) + v(s_i),$ | 1581.4 |

2. **Sensitivity analysis assessing the severity using individual-level covariates for the severity component following a Bernoulli distribution:**

To test the performance of our proposed method versus a joint method using the aggregated data to estimate the overall spatial distribution for the pattern and individual level data for severity following a Bernoulli distribution, we fitted additional models including age, sex, insurance scheme and minimum distance between severe cases as individual-level covariates for the severity component. To fit a joint hierarchical spatial model using the individual level data for the assessment of severity we constructed a model for the overall distribution of cases, specified as above in equation S.3 and other for the severe cases[2], where within each area we modeled the probability of case being severe or not, specified as follows:

$$q_{ij}|\eta_{ij}^{(2)} \sim Bernoulli(\eta_{ij}),$$ Equation S.7

$$Severity\ model: logit\left(\eta_{ij}^{(2)}\right) = \beta_0^{(2)} + \beta_1^{(2)} AGE(s_{ij}) + \beta_2^{(2)} SEX(s_{ij}) +$$

$$\beta_3^{(2)} INSURANCE(s_{ij}) + \beta_4^{(2)} DISTANCEKm(s_{ij}) + \beta_s f_s^i(s_i) + v(s_i),$$ Equation S.8

For the analysis of the severity in equation S.8 $q_{ij}$ is the severity status of a case (1 if the case is severe and 0 otherwise) in each neighborhood *i* which, conditional on the value of a second random field $\eta_{ij}^{(2)}$, follows a Bernoulli distribution, where $\eta_{ij}^{(2)}$ is the probability of an individual *j* being a severe case in neighborhood *i*. The logit$\left(\eta_{ij}^{(2)}\right) = log\left(\eta_{ij}^{(2)}/1 - \eta_{ij}^{(2)}\right)$ is the spatial field for the severity distribution at the individual level and the exponentiated $\beta^{(2)}$ coefficients are the odds ratio (OR) for severity: $\beta_0^{(2)}$is the intercept and the individual level fixed effects covariates for the severity included a categorical variable for age (under 15 years; 15 to 34 years; 35 to 54 years; and over 55 years): $AGE(s_{ij})$; an indicator variable for female sex: $SEX(s_{ij})$; the type of insurance: $INSURANCE(s_{ij})$ with 0 indicating subsidized scheme and 1 indicating a contributory scheme; and the minimum distance between severe cases per neighborhood $DISTANCEKm(s_{ij})$, which is the standardized nearest-neighbor (Euclidean) distance (km) between severe cases in each neighborhood. The logit of the probability of severity is decomposed as the sum of the fixed effects described above, a latent spatial effect $\beta_s f_s^i(s_i)$ that is proportional to the spatial latent effect in the log relative risk of overall dengue cases ($f_s^j(s_i)$), in equation (2) and an independent effect $v(s_i)$, which is a spatially unstructured random effect for the distribution of severe cases. The component $\beta_s f_s^i(s_i)$ in equation S.8 represents a single (common) random field, and includes a coefficient $\beta_s$ that makes the structured spatial effect for the severity ($\eta_{ij}^{(2)}$) proportional to the spatial effect of the pattern ( $\eta_i^{(1)}$) [2]; which is justified given that being a severe case is conditional on being a case in the first place.

**Table S6.** Posterior mean of the Odds Ratio and 95% credible intervals for covariates (fixed effects) on the **single-separated individual-level data model for severe dengue cases** in Medellin, 2013.

| | Besag (DIC =1416.05) | | | BYM (DIC=1415.26) | | | BYM2 (DIC=1415.29) | | |
|---|---|---|---|---|---|---|---|---|---|
| **Covariate** | **OR** | **2.5%** | **97.5%** | **OR** | **2.5%** | **97.5%** | **OR** | **2.5%** | **97.5%** |
| (Intercept) | 0.22 | 0.15 | 0.32 | 0.22 | 0.14 | 0.32 | 0.21 | 0.14 | 0.31 |
| Sex (Female) | 0.90 | 0.68 | 1.18 | 0.90 | 0.68 | 1.19 | 0.91 | 0.69 | 1.19 |
| Age (<15 years, Ref) | Ref. | - | - | Ref. | - | - | Ref. | - | - |
| 15-34 Years | 1.10 | 0.76 | 1.61 | 1.1 | 0.76 | 1.61 | 1.1 | 0.76 | 1.61 |
| 35-54 Years | 1.09 | 0.72 | 1.65 | 1.09 | 0.72 | 1.65 | 1.09 | 0.72 | 1.66 |
| >55 Years | 1.59 | 1.00 | 2.52 | 1.59 | 0.99 | 2.52 | 1.58 | 0.99 | 2.52 |
| Contributory Insurance | 0.83 | 0.63 | 1.11 | 0.83 | 0.62 | 1.11 | 0.82 | 0.61 | 1.09 |
| **Distance** to Severe cases (Km) | 0.45 | 0.23 | 0.83 | 0.45 | 0.24 | 0.84 | 0.47 | 0.24 | 0.90 |
| **Hyperparameter** | **Mean** | **2.5%** | **97.5%** | **Mean** | **2.5%** | **97.5%** | **Mean** | **2.5%** | **97.5%** |
| Precision for $f_s^j(s_i)$ | 18567.8 | 384.0 | 64604.6 | | | | 41.42 | 4.36 | 214.63 |
| Precision for $u_{(s_i)}$ | | | | 1837.3 | 118.0 | 6688.0 | | | |
| Precision for $f_s^j(s_i) + u_{(s_i)}$ | | | | 1768.7 | 105.2 | 6534.8 | | | |
| Phi for $u_{(s_i)}$ | | | | | | | 0.411 | 0.013 | 0.949 |

The joint model using individual data following a Bernoulli distribution for the assessment of the severity showed a slight decrease in the overall dengue rates by every 10% increase in the proportion of individuals with contributory scheme (SRR=0.93; 95%Cr.Int=0.89, 0.98), and a decrease overall dengue rate in low-SES (SRR=0.63; 95%Cr.Int=0.46, 0.85) and high-SES neighborhoods (SRR=0.64, 95%Cr.Int=0.43, 0.93) compared to medium SES neighborhoods. For severity, compared to people below 15 years old, severity tend to increase among people over 55 years old (OR=1.59; 95% Cr.Int=1.00, 2.53). The estimates for contributory insurance scheme showed high posterior uncertainty with 95% credible intervals covering the null value, (OR=0.84; 95%Cr.Int=0.63, 1.12). Female sex was not associated to overall dengue rates nor to severity. Estimates for distance indicated a decrease in the odds of severity for every kilometer increase in distance between severe cases within a neighborhood (OR=0.47; 95%Cr.Int=0.24, 0.87). The joint model using individual data for the severity showed smaller marginal variance of overall cases distribution (Variance= 0.99; 95%Cr.Int= 0.76, 1.32) and the Beta coefficient ($\beta_s$) for the spatial effect of severe cases indicated that after accounting for the other covariates in the model, the spatial distribution of severe dengue is not the same as the overall spatial distribution of dengue cases (OR= 1.09; 95% Cr.Int=0.90, 1.33) (Table S7). Although using a different likelihood for th3e assessment of severity (i.e., Bernoulli distribution) the Beta coefficient ($\beta_s$) for the spatial effect included the null value, this could be attributed to the sparce dataset for severity (n=247). Simulation exercises increasing the sample size showed that the mean spatial effect gain precision. Nonetheless, further research to identify the spatial dependencies and alternatives for modelling point process using individual location in infectious diseases is required.

**Table S7.** Posterior mean of the Standardized Rate Ratio (SRR), Odds Ratios (OR) and 95% credible intervals of covariates (fixed effects) for **the joint model for overall dengue distribution and individual-level data model for severity**.

| | Besag (DIC=2579.31) | | | BYM (DIC=2575.11) | | | BYM2 (DIC=2572.03) | | |
|---|---|---|---|---|---|---|---|---|---|
| **Pattern's Covariate** | **SRR** | **2.5%** | **97.5%** | **SRR** | **2.5%** | **97.5%** | **SRR** | **2.5%** | **97.5%** |
| Proportion of **female cases** | 1.01 | 0.97 | 1.05 | 1.0 | 0.96 | 1.04 | 1.0 | 0.96 | 1.05 |
| Proportion of **cases <20 years** old | 1.01 | 0.96 | 1.06 | 1.02 | 0.97 | 1.06 | 1.02 | 0.97 | 1.07 |
| Proportion of **contributory scheme** cases | 0.93 | 0.89 | 0.98 | 0.94 | 0.9 | 0.98 | 0.93 | 0.89 | 0.98 |
| **Medium** SES | Ref. | - | - | Ref. | - | - | Ref. | - | - |
| **Low** SES | 0.63 | 0.46 | 0.85 | 0.56 | 0.44 | 0.72 | 0.6 | 0.45 | 0.79 |
| **High** SES | 0.64 | 0.43 | 0.93 | 0.76 | 0.56 | 1.03 | 0.66 | 0.45 | 0.96 |
| Medium **Breteau Index** | 0.98 | 0.78 | 1.23 | 0.93 | 0.75 | 1.15 | 0.93 | 0.74 | 1.17 |
| **Severity Covariates** | **OR** | **2.5%** | **97.5%** | **OR** | **2.5%** | **97.5%** | **OR** | **2.5%** | **97.5%** |
| **Sex** (Female) | 0.9 | 0.68 | 1.19 | 0.9 | 0.68 | 1.18 | 0.9 | 0.68 | 1.19 |
| **Age (<**15 years, Ref) | Ref. | - | - | Ref. | - | - | Ref. | - | - |
| 15-34 Years | 1.09 | 0.76 | 1.59 | 1.09 | 0.75 | 1.58 | 1.09 | 0.75 | 1.59 |
| 35-54 Years | 1.09 | 0.72 | 1.65 | 1.09 | 0.72 | 1.65 | 1.09 | 0.72 | 1.65 |
| >55 Years | 1.59 | 1.0 | 2.53 | 1.6 | 1.0 | 2.55 | 1.59 | 1.0 | 2.53 |
| **Contributory** Insurance | 0.84 | 0.63 | 1.12 | 0.84 | 0.63 | 1.13 | 0.84 | 0.63 | 1.12 |
| **Distance** to Severe cases (Km) | 0.47 | 0.24 | 0.87 | 0.48 | 0.25 | 0.89 | 0.47 | 0.24 | 0.87 |
| **Hyperparameter** | **Mean** | **2.5%** | **97.5%** | **Mean** | **2.5%** | **97.5%** | **Mean** | **2.5%** | **97.5%** |
| Precision for $f_s^j(s_i)$ | 1.012 | 0.756 | 1.323 | | | | 1.998 | 1.51 | 2.597 |
| $\exp(\beta$ Coefficient for Severity for $g_s^j(s_{ij}))$ | 1.09 | 0.90 | 1.33 | 1.09 | 0.83 | 1.34 | 1.10 | 0.90 | 1.34 |
| Precision for $u_{(s_i)}$ | | | | 1.393 | 0.369 | 2.606 | | | |
| Precision for $f_s^j(s_i) + u_{(s_i)}$ | | | | 1858.3 | 97.99 | 8403.6 | | | |
| Phi for $u_{(s_i)}$ | | | | | | | 0.181 | 0.023 | 0.463 |

Proportion of woman, proportion of cases under 20 years old, and proportion of contributory scheme indicate 10% increase. Breteau Index reference group = Low; Age group reference= <15 years of age; Sex reference= Male; Health Insurance reference is Subsidized scheme. SES Level: Comparing cases in the Medium SES (Reference group) to Cases in the Low and High SES levels.

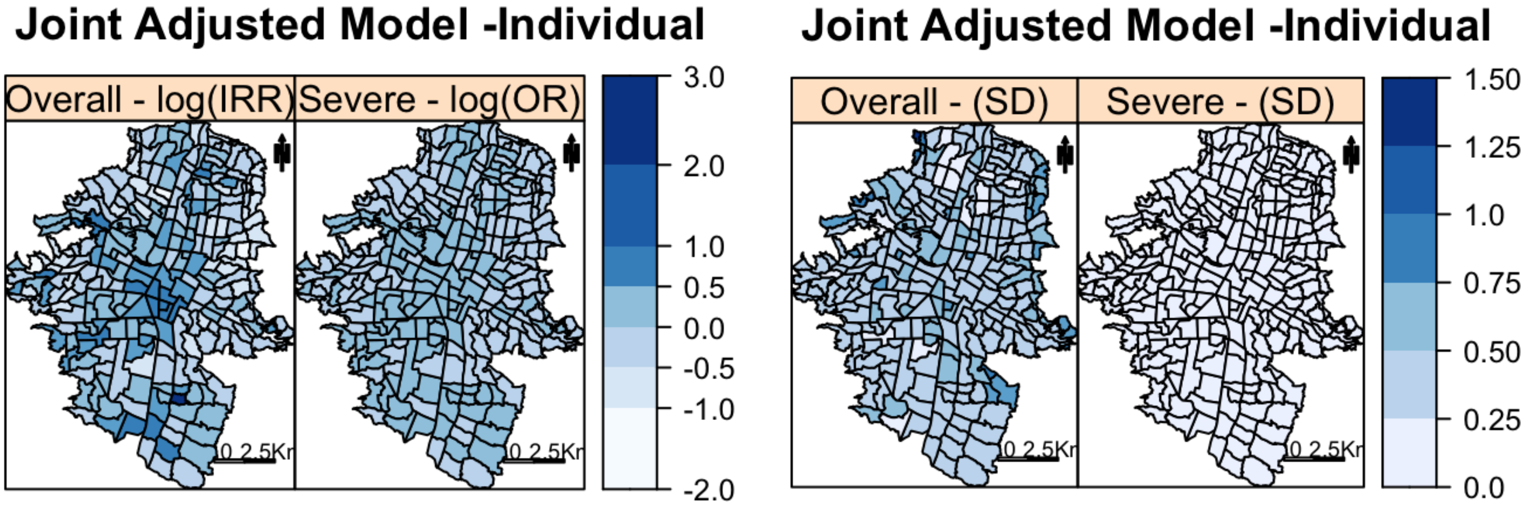


**Figure S4.** Estimated common spatial trend for overall dengue and severe dengue cases in Medellin, 2013. Neighborhood specific residual (Random Effects) Standardized Rate Ratio (nSRR) and Standard Deviation (SD). Neighborhood specific residual (Random Effects) Odds Ratio (nOR) and Standard Deviation (SD). Using individual-level data analysis for severity using a Bernoulli distribution and Besag parameterization.

Supplementary References:


1. Diggle PJ, Moraga P, Rowlingson B, Taylor BM. Spatial and Spatio-Temporal Log-Gaussian Cox Processes: Extending the Geostatistical Paradigm. *Statistical Science* 2013;**28**(4):542-563.
2. Illian JB, Martino S, Sorbye S, Gallego-Fernandez JB, Zunzunegui M, Esquivias MP, Travis JMJ. Fitting complex ecological point process models with Integrated Nested Laplace Approximation. *Methods in Ecology and Evolution* 2013;**4**:305-315.
3. Simpson D, Illian JB, Lindgren F, Sorbye S, Rue H. Going off grid: Computationally efficient inference for log-Gaussian Cox processes. *arXiv* 2017:1-26.
4. Illian JB, Sørbye SH, Rue H. A toolbox for fitting complex spatial point process models using integrated nested Laplace approximation (INLA). *The Annals of Applied Statistics* 2012:1499-1530.
5. Pinto Junior JA, Gamerman D, Paez MS, Alves Regina HF. Point pattern analysis with spatially varying covariate effects, applied to the study of cerebrovascular deaths. *Statistics in Medicine* 2015;**34**:1214-1226.
6. Waller LA, Gotway CA. Analyzing Public Health Data. . In: Shewhart WA, Wilks SS, eds. *Applied Spatial Statistics for Public Health Data*. Vol. 368 John Wiley & Sons, 2004;7-37.
7. Besag J, York J, Mollié A. Bayesian image restoration, with two applications in spatial statistics. *Annals of the Institute of Statistical Mathematics* 1991;**43**(1):1-20.
8. Blangiardo M, Cameletti M. Spatial modeling. *Spatial and Spatio-temporal Bayesian Models with R-INLA* John Wiley & Sons, Ltd, 2015;173-234.
9. Congdon P. Models for spatial outcomes and geographical association. *Applied Bayesian Modelling* John Wiley & Sons, Ltd, 2014;312-363.
10. Simpson D, Rue H, Riebler A, Martins TG, Sørbye SH. Penalising Model Component Complexity: A Principled, Practical Approach to Constructing Priors. *Statistical Science* 2017;**32**(1):1-28, 28.
